# Effect of virtual reality on upper limb recovery in early-stage stroke rehabilitation: A systematic review and meta-analysis

**DOI:** 10.64898/2025.12.11.25342076

**Authors:** Alba Hernández-Martínez, Manuel Fernandez-Escabias, Sofia Carrilho-Candeias, David Ruiz-González, Máriam Ramos-Teodoro, Mercedes Gil-Rodríguez, Laura del Olmo Iruela, Marta Rodríguez Camacho, Laura Amaya-Pascasio, Irene Pérez Ortega, Patricia Martínez-Sánchez, Alberto Soriano-Maldonado

**Author notes:** Corresponding author: Alba Hernández-Martínez.

## Abstract

**Background:** Virtual reality (VR) is increasingly used to enhance upper limb rehabilitation after stroke, yet its effectiveness during the acute and subacute phases—critical windows for neuroplasticity—remains unclear. This systematic review and meta-analysis evaluated the effects of VR-based interventions on upper limb outcomes in individuals within six months post-stroke and examined potential moderators of treatment response.

**Methods:** Five databases and grey literature were searched to June 2024. Randomised controlled trials including adults ≤6 months post-stroke and comparing VR-based rehabilitation with non-technological interventions were eligible. Random-effects models were used to pool mean differences (MDs). Subgroup analyses, random-effects meta-regressions, and sensitivity analyses were performed when data allowed. Risk of bias (RoB2) and certainty of evidence (GRADE) were assessed. The review was registered in PROSPERO (CRD420251104058).

**Results:** Twenty-eight trials (1,310 participants) were included. VR significantly improved motor impairment (FM-UE: MD = 3.82, 95% CI 2.16 5.48; I² = 83.8%) and movement speed (WMFT-time: MD = –0.52, 95% CI –0.87 to –0.18; I² = 0%). No significant effects were found for ARAT, BBT, or handgrip strength. Heterogeneity was substantial across most outcomes. Moderator analyses identified an age-related reduction in VR effects on handgrip strength and a time-dose interaction for BBT. Sensitivity analyses confirmed the stability of pooled estimates.

**Conclusions:** VR-based rehabilitation may enhance motor impairment and execution speed in the subacute phase after stroke, but evidence for functional improvements is limited. High heterogeneity, low methodological quality of included trials, and very low GRADE certainty for most outcomes warrant cautious interpretation. High-quality, standardised, and adequately powered trials are needed.

## Introduction

Stroke is a major global concern, currently ranking as the leading cause of disability and the second of mortality worldwide^1^. Its incidence is expected to double by 2050 due to population aging^2^. This trend places increasing demands on healthcare systems due to the long-term complications often associated with stroke^3^. Upper limb dysfunction is one of the most prevalent and disabling consequences, affecting approximately 80% of survivors^4^. It commonly impairs the ability to reach, grasp, and perform daily tasks^5^, with only 5-20% of patients fully recovering upper limb function^6^. Additionally, spasticity develops in up to half of cases, further limiting recovery^7^.

Rehabilitation plays a crucial role upper limb recovery after stroke^8^, relying on the brain’s capacity for neuroplasticity, the ability to reorganise motor outputs within surviving neural networks^9^. Evidence shows that neural activation in the lesioned hemisphere increases following structured upper limb rehabilitation programs^10^. Notably, most motor recovery occurs within the first months post-stroke, during the acute and subacute phases, which are considered critical windows for enhanced neuroplasticity^11^. To optimise recovery during this period, rehabilitation must be intense, repetitive, and engaging^11^. Studies suggest that meaningful improvements require hundreds of active repetitions per session to effectively stimulate neuroplastic mechanisms^12^. However, conventional therapy (CT) can often become monotonous and time-consuming, which may reduce patient motivation and adherence, ultimately limiting its effectiveness^13^. To address these limitations, technology-based approaches such as virtual reality (VR) have emerged as a promising tool in neurorehabilitation. VR provides interactive, computer-generated environments that replicate real-life or controlled settings in a safe, engaging, and cost-effective manner^14^. VR systems vary in immersion level, from non-immersive to fully immersive, based on the degree of sensory stimulation and interaction^15^. They may also differ in purpose, ranging from commercial gaming devices to stroke-specific rehabilitation tools, with different levels of gamification and therapeutic focus^16^. Beyond their adaptability, VR interventions may promote neuroplasticity by delivering high-repetition, task-specific, and patient-centred training aligned with key motor learning principles, such as intensity, salience, attentional focus, and performance feedback^17^. Moreover, VR may stimulate the mirror neuron system, enhancing recovery through action observation and imitation^18^.

Despite its growing popularity, the effectiveness of VR in stroke rehabilitation during the acute phase remains uncertain. Several systematic reviews and meta-analyses have reported positive effects on upper limb outcomes^19–23^. However, most of these combine patients across all stages of recovery or focus predominantly on the chronic phase, when the window for neuroplasticity has already diminished^11^. This limits our understanding of VR’s potential impact during the early post-stroke window, a critical period for recovery. Mixing recovery stages may also lead to misleading conclusions due to phenomena such as Simpson’s Paradox, where subgroup trends are obscured by aggregate data^24^. Additionally, some reviews aggregate heterogeneous outcome measures into broad categories of upper limb function, often using standardised mean differences, which can hinder clinical interpretation. Finally, only a few have explored whether treatment effects vary according to intervention characteristics (i.e., delivery format, VR system type, immersion level, dosage) or participant-related factors, such as age or sex distribution.

Therefore, the aim of this systematic review and meta-analysis was to evaluate the effectiveness of VR-based rehabilitation on upper limb motor outcomes, specifically in individuals with subacute stroke. This review also examined whether treatment effects varied according to key intervention and participant characteristics.

## Methods

### Search strategy and selection criteria

This systematic review and meta-analysis followed the PRISMA guidelines^25^ and was prospectively registered in the PROSPERO database (ID: CRD420251104058). Two reviewers (AHM and MFE) independently conducted a comprehensive literature search across PubMed, Scopus, Web of Science, and Cochrane, covering all records up to June 2024. Discrepancies were resolved through discussion with a third reviewer (SCC). The full search strategy is detailed in Supplementary Table 2. In addition, we manually searched the reference lists of relevant articles, previous reviews, and meta-analyses, as well as grey literature databases such as Google Scholar, TESEO, and OpenGrey.

Studies were eligible using the following criteria: (i) randomised controlled trial design; (ii) inclusion of adults who had experienced a stroke within the past six months (acute or subacute phase)^26^. Clinically, stroke refers to a sudden—or rapidly developing—neurological deficit of vascular origin, characterised by the abrupt onset of signs and symptoms that reflect dysfunction in specific brain regions^27^, diagnosed by imaging or neurological examination; (iii) implementation of a structured rehabilitation program based on VR technologies; VR was defined as “an advanced human-computer interface that allows realistic user interaction and immersion within a computer-generated environment”^28^; (iv) use of a comparison group receiving usual care or any non-VR intervention; and (v) assessment of at least one upper limb outcome measured before and after the intervention.

Studies were excluded if they met any of the following criteria: (i) The VR intervention was combined with robotic or electromechanical devices providing physical assistance to movement; (ii) The control group received any form of technology-based intervention involving virtual environments or interactive digital interfaces, in order to isolate the specific effects of VR-based rehabilitation compared to non-technological approaches; (iii) The intervention involved the use of pharmacological treatments; (iv) The post-stroke phase of participants was not reported; (v) The study employed studies robotic interventions without any screen display (i.e., without visual feedback); (vi) The sample included participants with mixed aetiologies (e.g., acquired brain injury) or mixed post-stroke phases (e.g., subacute and chronic), unless results were reported separately by pathology and by stroke-stroke phase. No additional restrictions were applied regarding stroke type (ischemic or haemorrhagic), lesion location (cortical or subcortical), or stroke severity.

For clarity, study eligibility was also defined according to the PICO framework: Population (P): adults in the acute or subacute phase of stroke recovery (≤6 months post-stroke), without restriction on stroke type, lesion location, or severity; Intervention (I): structured rehabilitation programs based on VR; Comparison (C): usual care or any non-technological rehabilitation approach; Outcome (O): Fugl-Meyer Upper Limb Assessment for Upper Extremity (FM-UE): is a validated and widely used scale to evaluate the upper limb motor impairment, focusing on joint movement, coordination, and reflexes in individuals recovering from stroke^29^; Action Research Arm Test (ARAT): assess functional arm use by measuring grasp, grip, pinch, and gross movement through goal-directed tasks relevant to daily activities^30^; Box and Block Test (BBT): quantifies gross manual dexterity by recording the number of blocks a person can grasp and transfer between compartments within 60 seconds using one hand^31^; Handgrip strength: typically assessed using a handheld dynamometer, reflects the maximal voluntary isometric force exerted by the forearm muscles and is widely used as an indicator of overall upper limb strength^32^; Wolf Motor Function Test (WMFT) Performance Time: assesses movement execution speed during timed tasks involving object manipulation, capturing both proximal and distal motor control necessary for everyday activities^33^.

## Data collection

Whenever available, the following information was extracted from each included study: first author, year of publication, country of corresponding author, study characteristics, participant demographics and baseline data, details of the intervention and comparator, methodological features, recruitment and retention rates, outcomes assessed and their measurement time points, data required for risk of bias assessments, and results. When multiple post-intervention assessments were reported, we extracted the data corresponding to the time point closest to the end of the intervention. In cases of missing data, study authors were contacted to obtain additional information. Review selection was managed using Rayyan software (Qatar Computing Research Institute, Qatar; www.rayyan.ai). In trials with more than two intervention arms, we combined the relevant intervention groups into a single composite group to avoid double counting the shared control group. Means and standard deviations were pooled following the formulas recommended in the Cochrane Handbook for Systematic Reviews of Interventions^34^.

This review is part of a broader project evaluating VR-based rehabilitation after stroke using a shared literature search and common eligibility criteria. The present review focuses on outcomes related to upper limb.

## Quality assessment

The Risk of Bias 2 (ROB2) tool was used to assess the methodological quality of the included studies^35^. Two independent reviewers (AHM, MFE) evaluated each study across the following domains: the randomisation process, deviation from intended interventions, missing outcome data, measurement of the outcome, selection of the reported result, and overall risk of bias. Each domain was rated as low risk, some concerns, or high risk of bias, following the Cochrane guidelines. Discrepancies were resolved through discussion or consultation with a third reviewer (SCC), when necessary.

## Strength of the body of evidence assessment (GRADE)

The certainty of the evidence for each primary outcome was evaluated using the Grading of Recommendations, Assessment, Development, and Evaluation (GRADE) approach^36^. Following standard procedures, the starting rating was set at “high,” but given that all included studies were randomised controlled trials, no downgrade was applied for study design. However, outcomes presenting serious or very serious limitations in specific domains—such as risk of bias, inconsistency, indirectness, imprecision, or publication bias—were downgraded accordingly by one or two levels. The GRADE ratings ranged from high to very low. Two reviewers (AHM and MFE) independently assessed each domain, and any disagreements were resolved through discussion with a third reviewer (SCC).

## Statistical analysis

A descriptive synthesis was conducted to summarise the characteristics and main findings of all included studies. When outcome data were sufficiently homogeneous in terms of measures of upper limb function, and at least three studies reported comparable outcomes, a quantitative synthesis was performed. Pooled effect estimates using mean differences (MDs) and corresponding 95% confidence intervals (CIs) were calculated using a random-effects meta-analysis, accounting for both within- and between-study variability. Statistical heterogeneity was assessed using the I² statistic.

When data allowed, subgroup analyses were conducted to explore potential sources of heterogeneity. Predefined subgroups included: (i) whether the intervention group performed VR alone or in combination with CT; (ii) whether the VR software used was specialised or commercially available; (iii) stroke phase, classified as early subacute (0–3 months post-stroke), late subacute (3–6 months post-stroke), or mixed (studies including participants within 0–6 months post-stroke)^26^; (iv) whether the total intervention time was matched between the experimental and control groups and (v) whether the VR software used was immersive or non-immersive. Initially, we also planned to conduct subgroup analyses comparing gamified versus non-gamified VR interventions; however, this information was not consistently reported across the included studies, and the analysis could not be performed.

To investigate potential moderators of treatment effect, random-effects meta-regression analyses were performed using the following study-level continuous variables: (i) difference in total intervention time between intervention and control groups; (ii) total duration of the intervention (weeks); (iii) total training time (minutes); (iv) mean participant age; and (v) percentage of female participants.

To explore potential publication bias and small-study effects, funnel plots were visually inspected, and Egger’s regression test was applied when ten or more studies were available for a given outcome^37^. Sensitivity analyses were performed using different estimators of the between-study variance and a leave-one-out approach, in which the meta-analysis was repeated after sequentially omitting each study, to assess the influence of individual studies on the pooled estimates.

## Results

A total of 7,932 records were initially identified through database searches. After removing duplicates and screening titles and abstracts, 491 full-text articles were assessed for eligibility. Ultimately, 28 randomised controlled trials published between 2014 and 2024 were included in the quantitative synthesis, comprising 1,310 participants (Figure 1). The included trials were conducted across diverse countries, with a predominance of studies from Asia and Europe. Mean participant age ranged from 46 to 74 years, and intervention durations ranged from two to eight weeks. Only four trials (14.3%) implemented immersive VR systems. Regarding the experimental conditions, most trials (n = 24) delivered VR in combination with CT, while four studies applied VR interventions alone. Control conditions also varied: 17 studies employed CT only, nine combined CT with additional activities, and two used other types of control interventions not based on usual care. Five primary outcomes related to upper limb motor recovery were analysed: FM-UE (n = 17), ARAT (n = 8), BBT (n = 10), and WMFT performance-time (n = 4) (Table S1).

**Figure 1.**
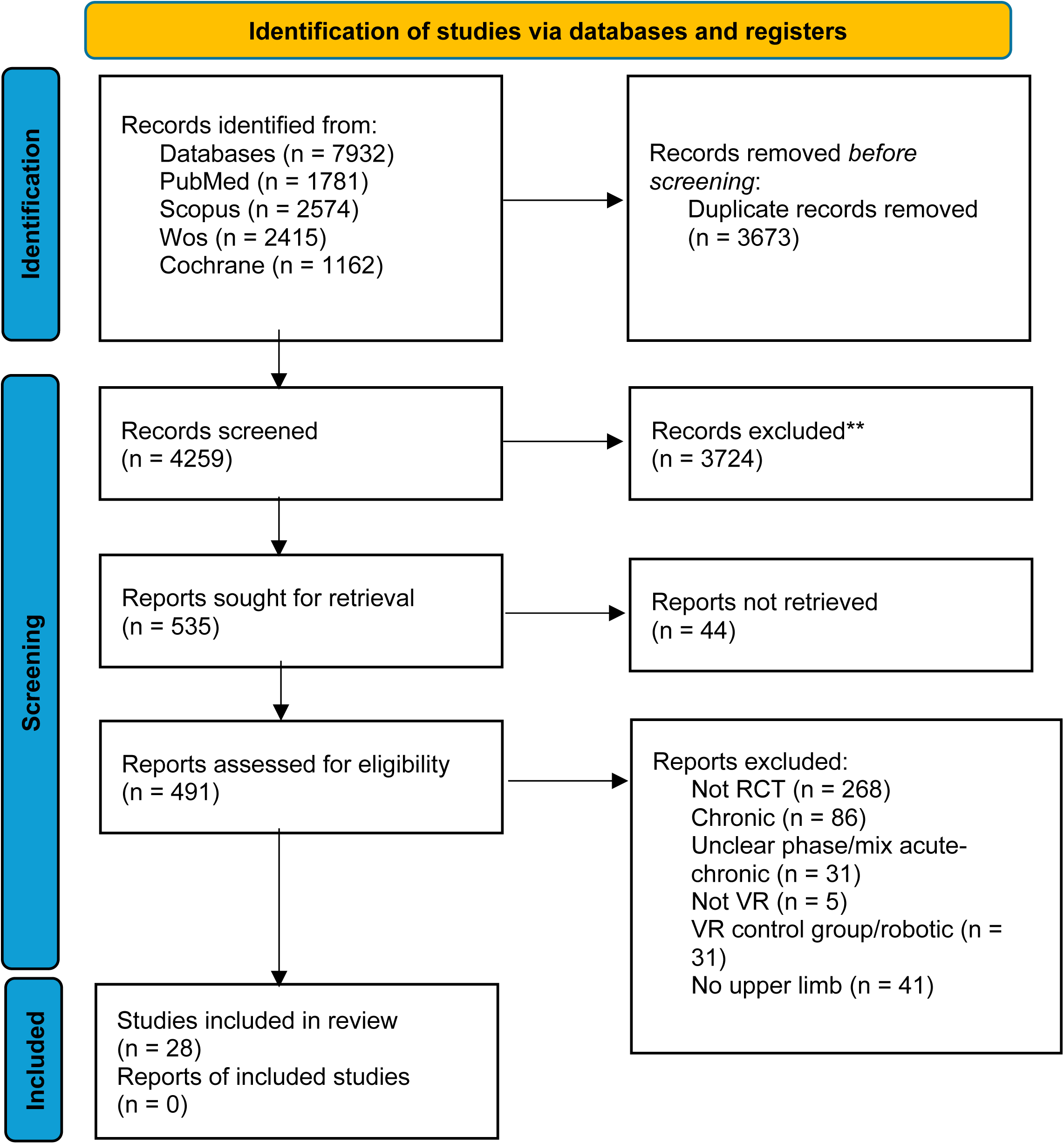
Flowchart of the search and selected studies

### Fugl Meyer Upper Limb Assessment for Upper Extremity (FM-UE)

Seventeen trials comprising 633 participants (35.8 % women) reported FM-UE and were included in this meta-analysis. In most studies (n=15), VR was delivered as a standalone intervention, while two trials combined VR with CT. Ten studies focused exclusively on participants in the early subacute phase, whereas the remaining seven included mixed phases (early and late). Only two trials employed immersive VR, and eight used specialised VR software. Total intervention time was matched between groups in all but three studies (Table S1). The pooled analysis revealed a statistically significant benefit of VR-based rehabilitation over control conditions for improving upper limb motor function with considerable heterogeneity (MD = 3.82; 95% CI: 2.16 to 5.48; I² = 83.8%). (Figure 3). No significant subgroup effects or moderator influences were identified in the meta-regression analyses (Supplementary Table S4, Figure S1, S2, S3, S4).

**Figure 2.**
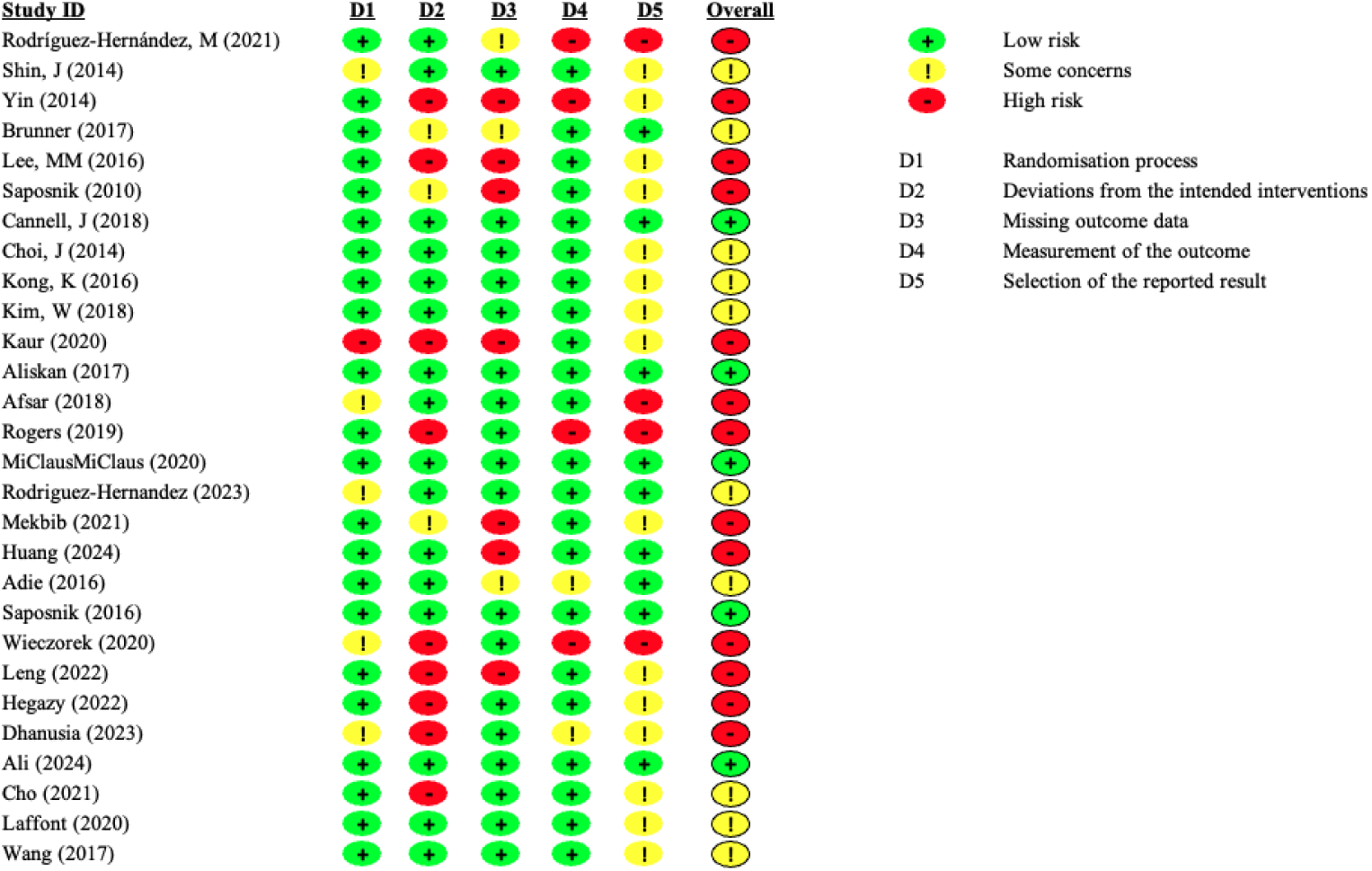
Methodological quality of the included studies assessed with the Risk of Bias 2 (RoB 2) tool

**Figure 3.**
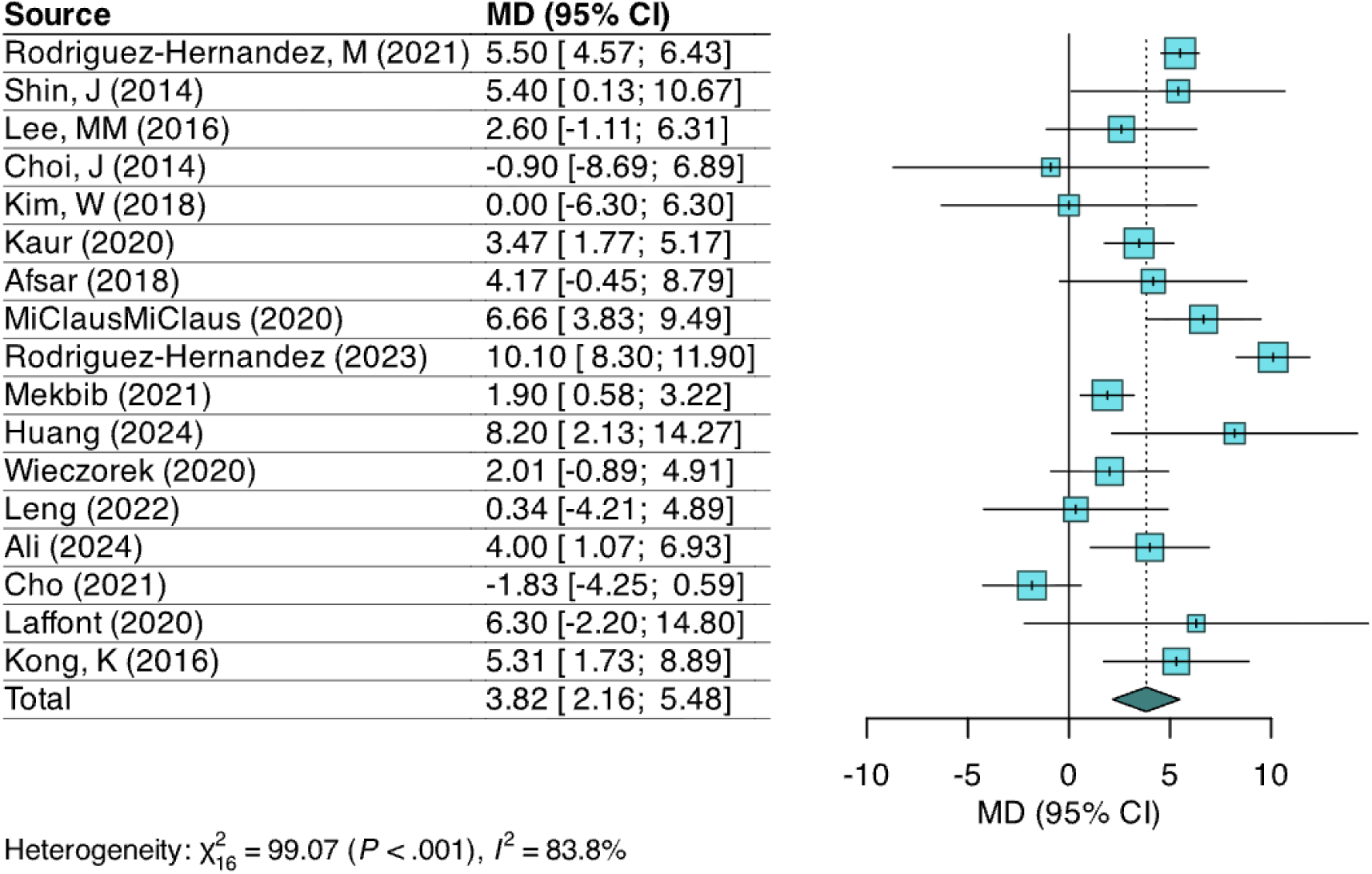
Forest plot of the effect of virtual reality-based interventions on upper-limb function measured with the Fugl-Meyer Assessment-Upper Extremity (FM-UE).

### Action Research Arm Test (ARAT)

Eight randomised controlled trials (n=645; 39.8% women) evaluated the effects of VR-based rehabilitation on upper limb function using the ARAT. In five studies, VR was combined with CT, whereas in the remaining three it was used as a standalone intervention. Five trials included participants in the early post-stroke phase (within 3 months), while the others enrolled mixed samples (≤ 6 months post-stroke). Only one study employed immersive VR technology. Half of the studies applied specialised VR software, while the other half used commercial gaming systems. In five studies, the total intervention time was matched between groups (Table S1). The pooled effect estimate showed a non-significant effect favouring VR over control with substantial heterogeneity (1.30; −4.19 to 6.79: I² = 96.9%) (Figure 4). No significant subgroup effects were observed (Figures S6, S7, S8). Although no significant, meta-regression analyses revealed a trend toward greater effects in studies with shorter intervention durations (weeks) (*β* = –2.16, 95% CI –4.50 to 0.19; *p* = 0.07; R^2^ = 59.09%) and smaller effects in those including a higher proportion of women participants (*β* = –0.40, 95% CI –0.89 to 0.09; *p* = 0.09; R² = 48.50%) (Table S5).

**Figure 4.**
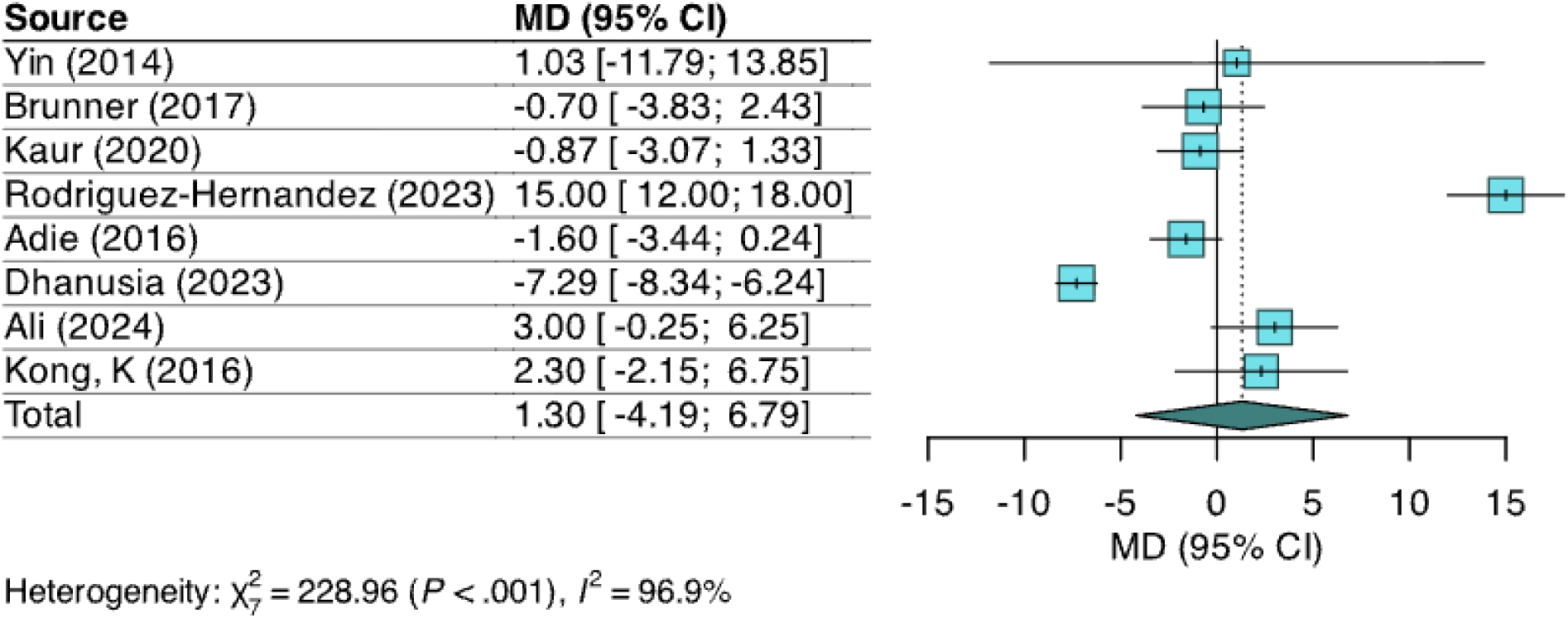
Forest plot of the effect of virtual reality-based interventions on upper-limb function measured with the Action Research Arm Test (ARAT).

### Box and Block Test (BBT)

Ten studies, including 484 participants (41.1% women), reported BBT. In all but one trial, VR was delivered as a standalone intervention. Most participants were in the early subacute phase, although two studies included mixed samples (early and late). None of the included trials used immersive VR, and four employed specialised software. In six studies, the total intervention time was matched between groups (Table S1). The pooled mean difference was 1.83 (−1.99 to 5.65; 80.6%), indicating a small, non-significant advantage for VR, with substantial heterogeneity (Figure 5). Subgroup analyses revealed a significant interaction between matched and unmatched intervention times (*p* = 0.005; Figure S5). Studies with unmatched total duration showed a non-significant but positive effect favouring VR (5.83, -1.12 to 12.78; 78.4%), whereas time matched trials demonstrated a small, non-significant effect in the opposite direction, favouring the control group (–1.51; –5.09 to 2.06; I² = 24.3%). This was supported by meta-regression, which showed a significant positive association between intervention time difference and treatment effect (*β* = 0.04, 95% CI 0.02 to 0.07; *p* = 0.007; R² = 78.1%), suggesting that larger time discrepancies were linked to greater effect sizes (Table S6). All remaining subgroup analyses and meta-regression models showed non-significant results (Supplementary Figure S9, Table S6).

**Figure 5.**
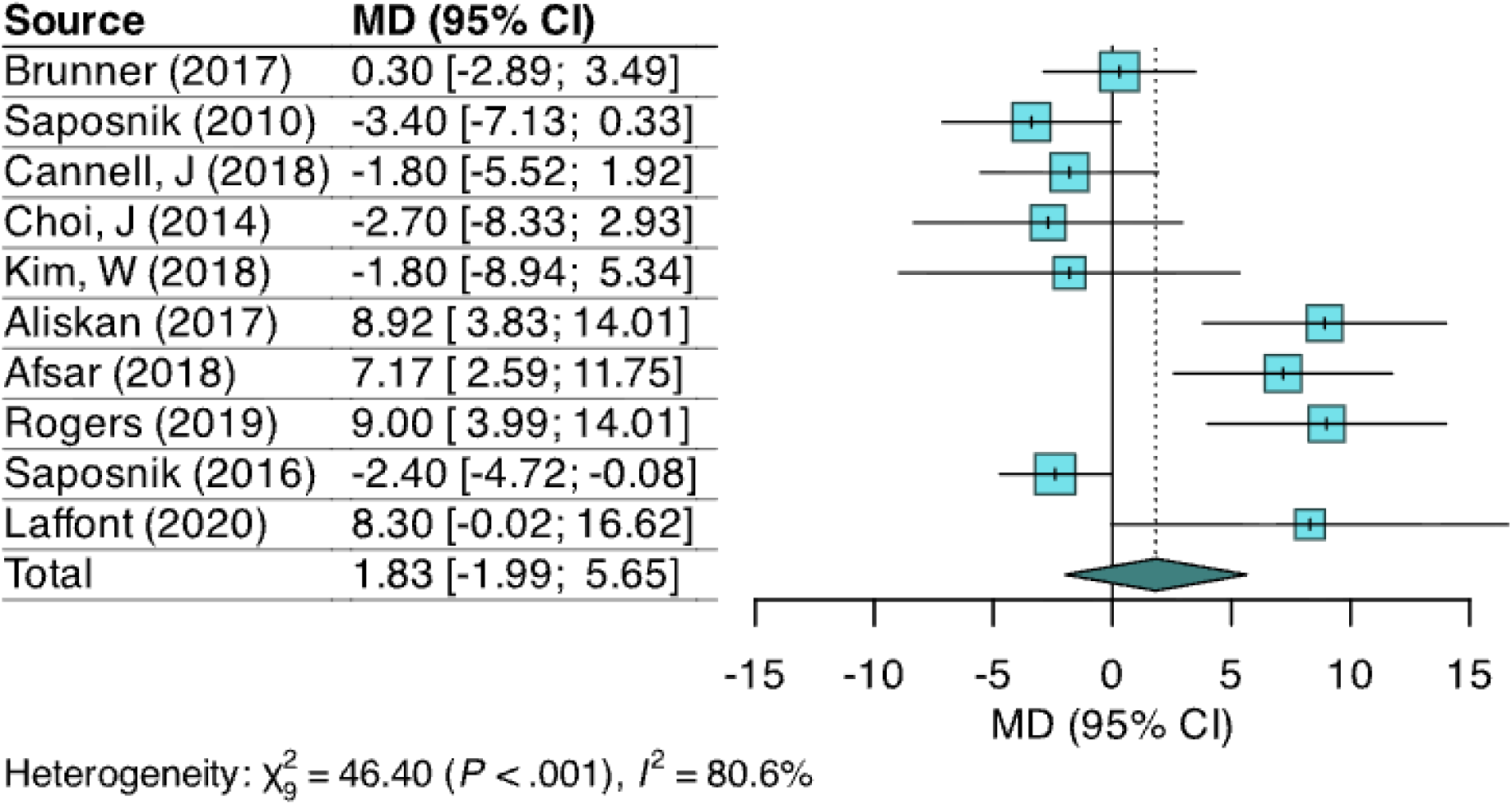
Forest plot of the effect of virtual reality-based interventions on upper-limb function measured with the Box and Block (BBT).

### Handgrip strength

This outcome was reported in six studies, including 217 participants (32.72% women). In four studies, VR was combined with CT, while the remaining two used VR alone. Most trials enrolled participants across both the early and late subacute phases, with only one study focusing exclusively on early subacute patients. Two studies implemented immersive VR systems, and none employed stroke-specialised software. Intervention time was matched between groups in all but one study (Table S1). The main pooled analysis revealed no significant effect of VR-based interventions on handgrip strength with considerable heterogeneity (0.15; −3.74 to 4.05; I² = 92.2%) (Figure 6). Meta-regression identified a significant inverse association between mean age and the treatment effect (*β* = −0.81, 95% CI −1.36 to −0.27; *p* = 0.017; R² = 91.0%), suggesting that younger participants benefited more from VR-based interventions (Table S7). No significant effects were found in the remaining subgroup (Figures S12, S13) or in the meta-regression analyses (Table S7).

**Figure 6.**
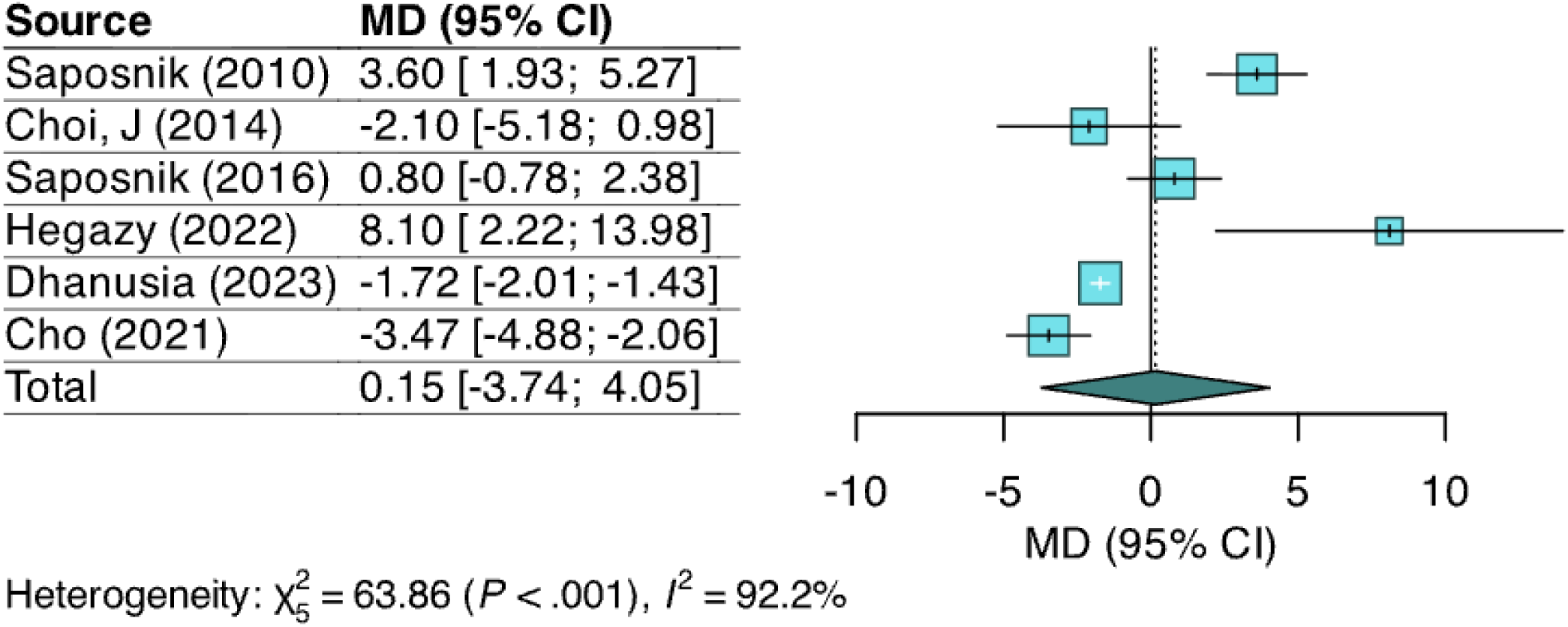
Forest plot of the effect of virtual reality-based interventions on handgrip strength.

### Wolf Motor Function Test (WMFT)-Performance time

In the present meta-analysis, we focused on the performance-time component, which quantifies the time (in seconds) needed to complete each task and serves as a sensitive indicator of motor execution efficiency. Data on WMFT ability scores were available in only two studies and were therefore excluded from the quantitative synthesis. The WMFT performance time was reported in four trials, including a total of 184 participants (34.78% women). In three studies, VR was combined with CT, while only one trial evaluated VR as a standalone intervention. Most participants were in the early subacute phase, and only one study used specialised VR software. None of the included studies employed immersive VR systems. The total intervention time was matched between groups in all but one study (Table S1). The pooled analysis showed a small but statistically significant benefit of VR-based rehabilitation on performance speed compared with control interventions with no observed heterogeneity (-0.52; −0.87 to −0.18; I² = 0%), (Figure 7). Due to the limited number of studies, subgroups and meta-regression analyses were not conducted.

**Figure 7.**
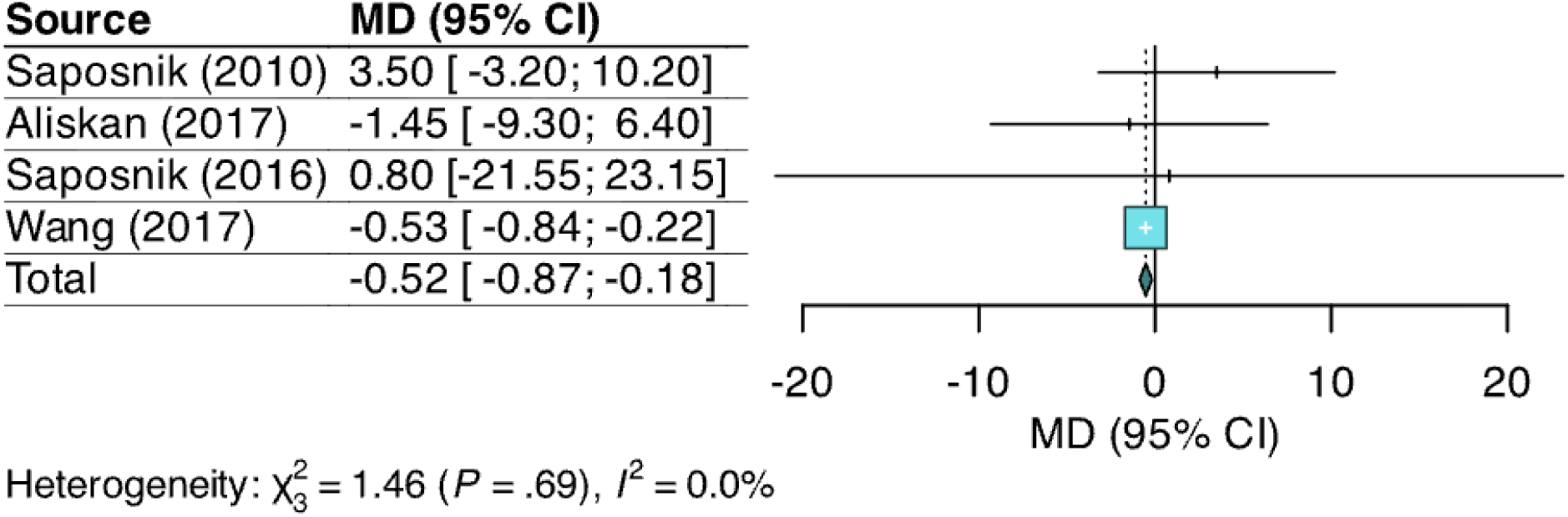
Forest plot of the effect of virtual reality-based interventions on upper-limb function measured with the Wolf Motor Function Test (WMFT) performance time.

## Methodological quality

The methodological quality of the included randomised controlled trials was evaluated using the Cochrane Risk of Bias 2 (RoB 2) tool (Figure 2). Only a few studies achieved an overall low risk of bias (n=5), while the majority were rated as having some concerns (n=11) or high risk (n=13), especially due to lack of blinding in outcome assessment and unclear pre-specification of analysis plans.

## GRADE

The GRADE approach was used to assess the certainty of the evidence across the primary outcomes. Table 1 summarises the overall ratings, considering risk of bias, inconsistency, indirectness, imprecision, and others. The certainty of evidence was rated as very low for FM-UE, ARAT, BBT, and handgrip strength, primarily due to serious or very serious concerns in methodological quality, high heterogeneity (I² > 75%), and the indirectness introduced by co-interventions or variability in comparators. Only WMFT-time achieved a moderate certainty rating, supported by consistent results and fewer concerns in the assessed domains. The most common reasons for downgrading were high risk of bias, particularly when a substantial proportion of studies showed high risk across ROB2 domains, and imprecision due to wide CI. No additional upgrading criteria were met, and no serious publication bias was detected through funnel plot inspection.

**Table 1.** Summary of findings: Effect of virtual reality–based rehabilitation on upper limb outcomes in patients with acute or subacute stroke

| Outcome | Studies | N. of patients | Certainty assessment |  |  |  |  | Absolute effect (95% CI) | Certainty |
| --- | --- | --- | --- | --- | --- | --- | --- | --- | --- |
|  |  |  | Risk of bias<sup>1</sup> | Inconsistency<sup>2</sup> | Indirectness<sup>3</sup> | Imprecision<sup>4</sup> | Other considerations |  |  |
| FM-UE | 17 | 633 | very serious | serious | serious | not serious | none | MD 3.95 higher (2.38 to 5.52) | ⊕○○○ VERY LOW |
| ARAT | 8 | 645 | very serious | serious | serious | serious | none* | MD 1.3 higher (4.19 to 6.79) | ⊕○○○ VERY LOW |
| BBT | 10 | 484 | serious | not serious | serious | serious | none | MD 1.83 higher (1.99 to 5.65) | ⊕○○○ VERY LOW |
| Handgrip | 6 | 217 | very serious | serious | serious | serious | none* | MD 0.15 higher (3.74 to 4.05) | ⊕○○○ VERY LOW |
| WMFT-time | 4 | 184 | serious | not serious | not serious | serious | none* | MD 0.52 lower (0.87 to 0.18) | ⊕⊕⊕○ MODERATE |
| **GRADE Working Group grades of evidence**<br>**High certainty:** The current evidence provides a very good indication of the likely effect, and the likelihood that the actual effect will be substantially different is low.<br>**Moderate certainty:** The current evidence provides a good indication of the likely effect, and the likelihood that the actual effect of the treatment will not be substantially different is moderate.<br>**Low certainty:** The current evidence provides some indication of the likely effect, but the likelihood that the actual effect will be substantially different is high.<br>**Very low certainty:** The current evidence does not provide a reliable indication of the likely effect, and the likelihood that the actual effect will be substantially different is very high. |  |  |  |  |  |  |  |  |  |
| FM-UE= Fugl-Meyer Assessment-Upper Extremity; ARAT= Action Research Arm Test; BBT=Box and block test; WMFT= Wolf Motor Function Test; CI=Confidence interval; MD=mean difference.<br><sup>1</sup> we downgraded by *serious* if over 50% of studies had some concerns or high risk, and by two levels *very serious* when high-risk studies represented a substantial proportion (≥40%) and no more than a minority were at low risk.<br><sup>2</sup> it was assessed using the I<sup>2</sup> statistic from the meta-analysis. Outcomes were rated as *serious* if I<sup>2</sup> > 75% and unexplained, and *not serious* if I<sup>2</sup> ≤ 40% or heterogeneity was clearly explained.<br><sup>3</sup> it was evaluated based on the variability of the comparators. Outcomes were rated as *serious* when a substantial proportion of studies used co-interventions or alternative therapies, and *not serious* when most studies used conventional therapy alone.<br><sup>4</sup> it was assessed based on the width and clinical relevance of the 95% confidence intervals. Outcomes were rated as *serious* when intervals were wide and crossed the line of no effect, and *not serious* when intervals were narrow and entirely on the side of benefit.<br>* publication bias assessment was not performed due to the small number of studies included and no upgrade (rating up) was given based on ‘large effect’, ‘plausible confounding’ and ‘dose response gradient’ domains. |  |  |  |  |  |  |  |  |  |

## Sensitivity analyses

Across all outcomes, the direction and statistical significance of the pooled estimates remained unchanged when individual studies were removed (Supplementary material). For ARAT, BBT, and handgrip strength, the pooled effects consistently remained non-significant, with very high heterogeneity persisting across all iterations. For FM-UE, the positive effect of VR was stable across all sensitivity analyses, with all leave-one-out models yielding statistically significant estimates. For WMFT, the limited number of studies precluded meaningful leave-one-out evaluation, but results were consistent across alternative variance estimators. Assessment of small-study effects was conducted only for outcomes with at least ten studies, as recommended. For FM-UE, the funnel plot showed no evidence of publication bias (Egger’s test: *p* = 0.501). Similarly, for BBT, no small-study effects were detected (Egger’s test: *p* = 0.103). Egger’s tests were not performed for the remaining outcomes due to the insufficient number of studies.

## Discussion

This systematic review and meta-analysis synthesised evidence from 28 randomised controlled trials evaluating the effectiveness of VR-based rehabilitation on upper limb recovery during the acute and subacute phases after stroke. Our findings suggests that VR interventions may significantly improve motor impairment (as measured by the FM-UE) and movement execution speed (WMF-performance), but do not appear to enhance functional arm use (ARAT), gross manual dexterity (BBT), or handgrip strength. These findings should be interpreted in the context of substantial heterogeneity, variations in intervention characteristics, and the limited use of immersive or stroke-specific systems.

Previous meta-analyses have examined the efficacy of VR-based rehabilitation after stroke, most of which pooled data across all recovery stages and outcomes, limiting their applicability to the subacute phase. In contrast, our review focuses specifically on the early post-stroke stage, a critical period of heightened neuroplasticity, and separates between distinct upper limb outcomes. Despite methodological differences, our findings of improvements in FM-UE are consistent with prior evidence. For instance, an umbrella review^38^ reported a larger pooled effect (SMD=2.10) favouring VR, without accounting for the stroke phase. A review of reviews^19^ concluded that VR interventions improve FM-UE scores regardless of dose or co-interventions, but most of the included studies involved chronic or mixed-phase patients. Regarding stroke phases, other systematic reviews have reported stronger effects in subacute versus chronic patients^6,20^. Nevertheless, it is essential to note that across most reviews, including ours, the average FM-UE improvements do not exceed the minimum clinically important difference of 9 points established for subacute patients^39^, suggesting that the observed benefits may have limited clinical relevance.

Although only four trials reported WMFT performance time, our analysis showed a statistically significant improvement in favour of VR, consistent with a prior meta-analysis^23^ that also reported positive effects, albeit based on just two trials. However, other reviews that did not stratify by stroke phase^19,21,40^ failed to find consistent benefits for this outcome. These discrepancies, likely due to heterogeneity in intervention design and stroke stage, underscore the need for more high-quality trials assessing VR’s impact on execution speed in the subacute phase.

In contrast, the lack of significant effects on functional outcomes such as ARAT^19,20^, BBT^19,20,23,40,41^ or handgrip strength^21,22^ has also been observed in prior reviews. Notably, most of these reviews were conducted in chronic stroke populations. A plausible explanation may lie in the different nature of the outcomes. FM-UE primarily measures motor impairment, including synergies, isolated movements, and basic coordination; whereas ARAT and BBT assess more complex tasks involving fine motor control, object manipulation, and functional use^29–31^. Many subacute VR interventions may emphasize repetitive practice of simple, gamified movements, which may primarily target impairment-level recovery without sufficient transfer to functional domains. Moreover, motor recovery trajectories vary considerably within the first six months post-stroke, with some patients experiencing spontaneous or proportional recovery limited to basic motor control^42^. Therefore, it is possible that the gains observed in motor impairment do not immediately translate into improvements in functional use, dexterity, or handgrip strength.

We were unable to explore the influence of immersion level in our analyses due to the limited number of trials using immersive systems. However, another study has reported a dose-dependent gradient of effectiveness: immersive VR tends to produce greater gains in FM-UE than non-immersive or commercial systems^43^. Similarly, another review^20^ has shown that immersive VR systems improve FM-UE (MD=3.04) and BBT (MD=2.85), though no consistent effects were found for ARAT. Although these findings are based on studies in patients with chronic stroke, they suggest that the degree of immersion may play a relevant role in motor recovery, and future studies should further explore this variable using standardised protocols and immersive technologies tailored to stroke rehabilitation during the acute/subacute phase.

Regarding intervention dosage, our analyses revealed a trend (though not statistically significant) suggesting that shorter intervention durations were associated with greater ARAT improvements. For BBT, trials with unmatched intervention time between groups showed larger effects favouring VR, and greater between-groups differences in intervention time were positively associated with effect sizes. No time effects were observed for the other outcomes. These patterns align with previous reviews reporting stronger effects with longer intervention time (>15h)^22^, especially for FM-UE or BBT^21,41^. Interestingly, shorter VR sessions may be more beneficial for FM-UE^23^, and that dose-response relationships may be more pronounced in chronic patients than in subacute ones^20^.

Among participant-level moderators, only age showed a significant association: each additional year was associated with a decrease of 0.9 kg in the effect of VR on handgrip strength. This result is plausible given the relationship between age and muscle strength^44^ and has also been observed in previous studies on FM-UE outcomes^23^. Regarding sex, a non-significant trend was observed, indicating that studies with a higher proportion of women reported smaller ARAT effects. Given the male predominance in most samples, further research is warranted to examine potential sex differences in VR responsiveness.

As for the type of intervention (stroke-specific vs. commercial) and delivery format (VR alone vs. in combination with CT), comparisons were only feasible for a limited number of outcomes and showed no significant results. Most included studies used non-specific commercial systems and combined VR with usual care, but the definition of CT varied considerably across trials, limiting meaningful comparisons. The limited use of immersive or stroke-specific systems may partially explain the lack of functional gains, as these features are essential for promoting task salience and real-life transfer^45^.

From a clinical perspective, our findings offer useful insight but should be interpreted with caution. While VR-based rehabilitation appears to support improvements in motor impairment and execution speed, the size of these changes may not be large enough to produce meaningful functional gains in everyday life. This is especially relevant for FM-UE, where the average improvements fell below the threshold typically considered clinically important in subacute stroke. Additionally, the overall quality of the evidence was low for most outcomes, mainly due to methodological limitations, high variability between studies, and imprecise results. These factors suggest that, although VR shows promise as a complementary rehabilitation tool, its benefits may vary depending on the type of intervention, patient profile, and how it is integrated into broader therapy programs.

This review has some strengths, and they include its exclusive focus on the early post-stroke recovery phase, underrepresented in the literature despite being critical for neuroplasticity changes, and its outcome-specific approach using MD, allowing better clinical interpretation. We also explored several potential moderators to explain variability in results. However, several limitations should be noted. First, most analyses exhibited substantial heterogeneity, and the methodological quality of the included trials was generally low, with many studies showing unclear or high risk of bias across multiple domains. This reduces confidence in the pooled estimates and underscores the need for more rigorous trial designs in this research area. Additionally, the small number of studies available for some outcomes limited statistical power for subgroup analyses, meta-regression analyses, and assessments of publication bias. Egger’s tests were conducted only for FM-UE and BBT, as these were the only outcomes with at least ten studies; for all other outcomes, publication bias could not be formally evaluated. Finally, several trials included very small samples and limited representation of women, highlighting the need for larger, better-powered, and more balanced studies in this population.

Future research should prioritise the development of high-quality randomised controlled trials focusing specifically on the acute and subacute phases of stroke recovery. Trials should incorporate immersive and stroke-specific VR systems, ensure adequate control of intervention dosage, and standardise comparison groups to enhance interpretability across studies. In addition, future work should consider sex-balanced samples and include subgroup analyses to explore differential responses by age, sex, or stroke severity. There is also a need to assess long-term functional outcomes and the transfer of motor gains into meaningful daily activities.

## Conclusion

In summary, this meta-analysis suggests that virtual reality-based rehabilitation may improve motor impairment and movement speed in the early stages after stroke, particularly as reflected in FM-UE and WMFT performance-time outcomes. However, no significant effects were observed for functional use, dexterity, or strength measures such as ARAT, BBT, or handgrip. While these findings highlight the potential of VR as a complementary tool to conventional therapy, they also underscore important limitations, including substantial heterogeneity, high risk of bias in many studies, and very low certainty of the evidence for most outcomes according to GRADE. Future studies should prioritise standardised, well-powered trials with clearer intervention protocols, improved methodological quality, and sex-balanced representation of participant characteristics to better understand which individuals benefit most from VR and under what conditions.

## Data Availability

All data included in this study were extracted from published articles. No new datasets were generated. All extracted data used for analyses are available within the manuscript and its supplementary material.

## References

[1] World Stroke Organization. Stroke Advocacy Brochure.

[2] Anderson CS. The global burden of stroke : persistent and disabling The global burden of neurological disorders. Lancet Neurol. 2019;18(5):417–418. doi:10.1016/S1474-4422(19)30030-4

[3] Avan A, Digaleh H, Napoli M Di, Stranges S, Behrouz R, Shojaeianbabaei G. Socioeconomic status and stroke incidence , prevalence , mortality , and worldwide burden : an ecological analysis from the Global Burden of Disease Study 2017. Published online 2019.

[4] Ahmed N, Mauad VAQ, Gomez-Rojas O, et al. The Impact of Rehabilitation-oriented Virtual Reality Device in Patients With Ischemic Stroke in the Early Subacute Recovery Phase: Study Protocol for a Phase III, Single-Blinded, Randomized, Controlled Clinical Trial. J Cent Nerv Syst Dis. 2020;12. doi:10.1177/1179573519899471

[5] World Health Organization. World Health Organization. Published 2025. https://www.who.int/health-topics/cardiovascular-diseases/#tab=tab_1

[6] Soleimani M, Ghazisaeedi M, Heydari S. The efficacy of virtual reality for upper limb rehabilitation in stroke patients: a systematic review and meta-analysis. BMC Med Inform Decis Mak. 2024;24(1):135. doi:10.1186/s12911-024-02534-y

[7] Lackritz H, Parmet Y, Frenkel-Toledo S, et al. Effect of post-stroke spasticity on voluntary movement of the upper limb. J Neuroeng Rehabil. 2021;18(1):1–14. doi:10.1186/s12984-021-00876-6

[8] Hatem SM, Saussez G, della Faille M, et al. Rehabilitation of motor function after stroke: A multiple systematic review focused on techniques to stimulate upper extremity recovery. Front Hum Neurosci. 2016;10(SEP2016):1–22. doi:10.3389/fnhum.2016.00442

[9] Hordacre B, Austin D, Brown KE, et al. Evidence for a Window of Enhanced Plasticity in the Human Motor Cortex Following Ischemic Stroke. Neurorehabil Neural Repair. 2021;35(4):307–320. doi:10.1177/1545968321992330

[10] Richards LG, Stewart KC, Woodbury ML, Senesac C, Cauraugh JH. Movement-dependent stroke recovery: A systematic review and meta-analysis of TMS and fMRI evidence. Neuropsychologia. 2008;46(1):3–11. doi:10.1016/j.neuropsychologia.2007.08.013

[11] Langhorne P, Bernhardt J, Kwakkel G. Stroke rehabilitation. Lancet. 2011;377(9778):1693-1702. doi:10.1016/S0140-6736(11)60325-5

[12] Birkenmeier RL, Prager EM, Lang CE. Translating animal doses of task-specific training to people with chronic stroke in 1-hour therapy sessions: A proof-of-concept study. Neurorehabil Neural Repair. 2010;24(7):620–635. doi:10.1177/1545968310361957

[13] Li W, Zhu G, Lu Y, et al. The relationship between rehabilitation motivation and upper limb motor function in stroke patients. Front Neurol . 2024;15(May):1–12. doi:10.3389/fneur.2024.1390811

[14] Weiss PL, Kizony R, Feintuch U, Rand D, Katz N. Textbook of Neural Repair and Rehabilitation. Cambridge University Press; 2006.

[15] Slater M. Immersion and the illusion of presence in virtual reality. Br J Psychol. 2018;109(3):431–433. doi:10.1111/bjop.12305

[16] Rodríguez-Hernández M, Polonio-López B, Corregidor-Sánchez AI, Martín-Conty JL, Mohedano-Moriano A, Criado-Álvarez JJ. Can specific virtual reality combined with conventional rehabilitation improve poststroke hand motor function? A randomized clinical trial. J Neuroeng Rehabil. 2023;20(1):1–14. doi:10.1186/s12984-023-01170-3

[17] Verschure PFMJ. Neuroscience, virtual reality and neurorehabilitation: Brain repair as a validation of brain theory. Proc Annu Int Conf IEEE Eng Med Biol Soc EMBS. Published online 2011:2254–2257. doi:10.1109/IEMBS.2011.6090428

[18] Da Silva Cameiro M, Bermúdez I Badia S, Duarte E, Verschure PFMJ. Virtual reality based rehabilitation speeds up functional recovery of the upper extremities after stroke: A randomized controlled pilot study in the acute phase of stroke using the Rehabilitation Gaming System. Restor Neurol Neurosci. 2011;29(5):287–298. doi:10.3233/RNN-2011-0599

[19] Bargeri S, Scalea S, Agosta F, et al. Effectiveness and safety of virtual reality rehabilitation after stroke: an overview of systematic reviews. eClinicalMedicine. 2023;64:1–15. doi:10.1016/j.eclinm.2023.102220

[20] Kenea CD, Abessa TG, Lamba D. Immersive Virtual Reality in Stroke Rehabilitation : A Systematic Review and Meta-Analysis of Its Efficacy in Upper Limb Recovery. Published online 2025.

[21] Chen J, Or CK, Chen T. Effectiveness of Using Virtual Reality–Supported Exercise Therapy for Upper Extremity Motor Rehabilitation in Patients with Stroke: Systematic Review and Meta-analysis of Randomized Controlled Trials. J Med Internet Res. 2022;24(6):1–26. doi:10.2196/24111

[22] Laver KE, Lange B, George S, et al. Virtual reality for stroke rehabilitation. Cochrane Database Syst Rev. 2025;2025(6). doi:10.1002/14651858.CD008349.pub5

[23] Xu S, Xu Y, Wen R, Wang J, Qiu Y, Chan CCH. Virtual Reality Enhanced Exercise Training in Upper Limb Function of Patients With Stroke : Meta-Analytic Study Corresponding Author : 2025;27. doi:10.2196/66802

[24] Altman DG, Deeks JJ. Meta-analysis , Simpson ’ s paradox , and the number needed to treat. 2002;5:1–5.

[25] Page MJ, Moher D, Bossuyt PM, et al. PRISMA 2020 explanation and elaboration: Updated guidance and exemplars for reporting systematic reviews. BMJ. 2021;372. doi:10.1136/bmj.n160

[26] Bernhardt J, Hayward KS, Kwakkel G, et al. Agreed definitions and a shared vision for new standards in stroke recovery research: The Stroke Recovery and Rehabilitation Roundtable taskforce. Int J Stroke. 2017;12(5):444–450. doi:10.1177/1747493017711816

[27] WHO Task Force on Stroke and Other Cerebrovascular Disorders. Special Report From the World Health Organization Stroke *—* 1989.; 1989. doi:10.1161/01.STR.20.10.1407

[28] Laver KE, Lange B, George S, Deutsch JE, Saposnik G, Crotty M. Virtual reality for stroke rehabilitation. Cochrane Database Syst Rev. 2017;2018(1):CD008349. doi:10.1002/14651858.CD008349.pub4

[29] Gladstone DJ, Danells CJ, Black SE. The Fugl-Meyer Assessment of Motor Recovery after Stroke: A Critical Review of Its Measurement Properties. Published online 2002.

[30] Pike S, Lannin NA, Wales K, Cusick A. Special Issue Article A systematic review of the psychometric properties of the Action Research Arm Test in neurorehabilitation. 2018;(August):449–471. doi:10.1111/1440-1630.12527

[31] Julien M, Amours JD, Leduc M, et al. Physical & Occupational Therapy In Geriatrics Responsiveness of the Box and Block Test with Older Adults in Rehabilitation Responsiveness of the Box and Block Test with Older Adults. 2017;3181(November). doi:10.1080/02703181.2017.1356897

[32] Williams ER, Wilson HK, Ross RE, et al. Relative handgrip strength as a vitality measure in US stroke survivors. Disabil Rehabil. 2024;46(26):6345–6351. doi:10.1080/09638288.2024.2327488

[33] Wolf SL, Catlin PA, Ellis M, Archer AL, Morgan B, Piacentino A. Assessing Wolf Motor Function Test as Outcome Measure for Research in Patients After Stroke. Published online 2001:1635–1639. doi:10.1161/01.STR.32.7.1635

[34] Axon E, Dwan K, Richardson R. Multiarm studies and how to handle them in a meta - analysis : A tutorial. 2023;(November):1–6. doi:10.1002/cesm.12033

[35] Sterne JAC, Savović J, Page MJ, et al. RoB 2: A revised tool for assessing risk of bias in randomised trials. BMJ. 2019;366:1–8. doi:10.1136/bmj.l4898

[36] Schünemann H, Brożek J, Guyatt G O. GRADE handbook for grading quality of evidence and strength of recommendations. Published 2013. Accessed April 28, 2022. https://gdt.gradepro.org/app/handbook/handbook.html

[37] Pustejovsky JE, Rodgers MA. Testing for funnel plot asymmetry of standardized mean differences. 2019;(June 2018):57–71. doi:10.1002/jrsm.1332

[38] Wu J, Zeng A, Chen Z, et al. Effects of virtual reality training on upper limb function and balance in stroke patients: Systematic review and meta-meta-analysis. J Med Internet Res. 2021;23(10). doi:10.2196/31051

[39] Arya KN, Verma R, Garg RK. Estimating the Minimal Clinically Important Difference of an Upper Extremity Recovery Measure in Subacute Stroke Patients. 2016;9357(March). doi:10.1310/tsr18s01-599

[40] Leong SC, Tang YM, Toh FM, Fong KNK. Examining the effectiveness of virtual, augmented, and mixed reality (VAMR) therapy for upper limb recovery and activities of daily living in stroke patients: a systematic review and meta-analysis. J Neuroeng Rehabil. 2022;19(1):93. doi:10.1186/S12984-022-01071-X

[41] Mekbib DB, Han J, Zhang L, et al. Virtual reality therapy for upper limb rehabilitation in patients with stroke: a meta-analysis of randomized clinical trials. Brain Inj. 2020;34(4):456–465. doi:10.1080/02699052.2020.1725126

[42] Vliet R Van Der, Selles RW, Andrinopoulou E, et al. Recovery after Stroke : A Mixture Model. Published online 2020:383–393. doi:10.1002/ana.25679

[43] Hao J, He Z, Yu X, Remis A. Comparison of immersive and non-immersive virtual reality for upper extremity functional recovery in patients with stroke: a systematic review and network meta-analysis. Neurol Sci. 2023;44(8):2679–2697. doi:10.1007/s10072-023-06742-8

[44] McLeod M, Breen L, Hamilton DL, Philp A. Live strong and prosper: the importance of skeletal muscle strength for healthy ageing. Biogerontology. 2016;17(3):497–510. doi:10.1007/s10522-015-9631-7

[45] Donati D, Pinotti E, Mantovani M, et al. The Role of Immersive Virtual Reality in Upper Limb Rehabilitation for Subacute Stroke : A Review. Published online 2025:1–21.

