## Supplementary material for "Effect of virtual reality on upper limb recovery in early-stage stroke rehabilitation: A systematic review and meta-analysis": SUPPLEMENTARY_preprint.pdf


**Authors:**

Alba Hernández-Martínez[1], Manuel Fernandez-Escabias[2], Sofia Carrilho-Candeias[2], David Ruiz-González[1,3], Máriam Ramos-Teodoro[1,4], Mercedes Gil-Rodríguez[5], Laura del Olmo Iruela[6], Marta Rodríguez Camacho[7], Laura Amaya-Pascasio[7], Irene Pérez Ortega[8], Patricia Martínez-Sánchez[7,9], Alberto Soriano-Maldonado[1,3]

**Affiliations:**

1. SPORT Research Group (CTS-1024), CIBIS (Centro de Investigación para el Bienestar y la Inclusión Social) Research Center), University of Almería, Almería, Spain.
2. Department of Physiology, Faculty of Medicine, University of Granada, Granada, Spain.
3. Department of Education, Faculty of Education Sciences, University of Almería, Almería, Spain.
4. Department of Nursing, Physiotherapy and Medicine, University of Almería, Almería, Spain.
5. Fundación para la Investigación Biosanitaria de Andalucía Oriental (FIBAO), Spain.
6. Department of Physical Medicine and Rehabilitation, San Cecilio University Hospital, Granada, Spain.
7. Department of Neurology, Torrecárdenas University Hospital, Almería, Spain.
8. Department of Neurology, Virgen de las Nieves University Hospital, Granada, Spain.
9. Faculty of Health Sciences, Health Research Centre (CEINSA), University of Almería, Spain.



**Funding:**

This work was partially supported by the RESET Project (Public–Private Collaboration Project, ref. CPP2021-008497), funded by MCIN/AEI/10.13039/501100011033 and by the European Union NextGenerationEU/PRTR.


**Conflicts of interest:**

The authors declare no competing interests.

### PRISMA 2020 Checklist

| Section and Topic | Item # | Checklist item | Location where item is reported |
|---|---|---|---|
| **TITLE** | | | |
| Title | 1 | Identify the report as a systematic review. | 1 |
| **ABSTRACT** | | | |
| Abstract | 2 | See the PRISMA 2020 for Abstracts checklist. | 2 |
| **INTRODUCTION** | | | |
| Rationale | 3 | Describe the rationale for the review in the context of existing knowledge. | 2-3 |
| Objectives | 4 | Provide an explicit statement of the objective(s) or question(s) the review addresses. | 3 |
| **METHODS** | | | |
| Eligibility criteria | 5 | Specify the inclusion and exclusion criteria for the review and how studies were grouped for the syntheses. | 4 |
| Information sources | 6 | Specify all databases, registers, websites, organisations, reference lists and other sources searched or consulted to identify studies. Specify the date when each source was last searched or consulted. | 4 |
| Search strategy | 7 | Present the full search strategies for all databases, registers and websites, including any filters and limits used. | Supplementary table S3 |
| Selection process | 8 | Specify the methods used to decide whether a study met the inclusion criteria of the review, including how many reviewers screened each record and each report retrieved, whether they worked independently, and if applicable, details of automation tools used in the process. | 4 |
| Data collection process | 9 | Specify the methods used to collect data from reports, including how many reviewers collected data from each report, whether they worked independently, any processes for obtaining or confirming data from study investigators, and if applicable, details of automation tools used in the process. | 5 |
| Data items | 10a | List and define all outcomes for which data were sought. Specify whether all results that were compatible with each outcome domain in each study were sought (e.g. for all measures, time points, analyses), and if not, the methods used to decide which results to collect. | 5 |
| | 10b | List and define all other variables for which data were sought (e.g. participant and intervention characteristics, funding sources). Describe any assumptions made about any missing or unclear information. | 5 |
| Study risk of bias assessment | 11 | Specify the methods used to assess risk of bias in the included studies, including details of the tool(s) used, how many reviewers assessed each study and whether they worked independently, and if applicable, details of automation tools used in the process. | 6 |
| Effect measures | 12 | Specify for each outcome the effect measure(s) (e.g. risk ratio, mean difference) used in the synthesis or presentation of results. | 6 |
| Synthesis methods | 13a | Describe the processes used to decide which studies were eligible for each synthesis (e.g. tabulating the study intervention characteristics and comparing against the planned groups for each synthesis (item #5)). | 6-7 |
| | 13b | Describe any methods required to prepare the data for presentation or synthesis, such as handling of missing summary statistics, or data conversions. | 7 |
| | 13c | Describe any methods used to tabulate or visually display results of individual studies and syntheses. | 7 |
| | 13d | Describe any methods used to synthesize results and provide a rationale for the choice(s). If meta-analysis was performed, describe the model(s), method(s) to identify the presence and extent of statistical heterogeneity, and software package(s) used. | 6-7 |
| | 13e | Describe any methods used to explore possible causes of heterogeneity among study results (e.g. subgroup analysis, meta-regression). | 7 |
| | 13f | Describe any sensitivity analyses conducted to assess robustness of the synthesized results. | 7 |
| Reporting bias assessment | 14 | Describe any methods used to assess risk of bias due to missing results in a synthesis (arising from reporting biases). | 7 |
| Certainty | 15 | Describe any methods used to assess certainty (or confidence) in the body of evidence for an outcome. | 6 |

| Section and Topic | Item # | Checklist item | Location where item is reported |
|---|---|---|---|
| assessment | | | |
| **RESULTS** | | | |
| Study selection | 16a | Describe the results of the search and selection process, from the number of records identified in the search to the number of studies included in the review, ideally using a flow diagram. | Figure 1 |
| | 16b | Cite studies that might appear to meet the inclusion criteria, but which were excluded, and explain why they were excluded. | NA |
| Study characteristics | 17 | Cite each included study and present its characteristics. | Table S1 |
| Risk of bias in studies | 18 | Present assessments of risk of bias for each included study. | Figure 2 |
| Results of individual studies | 19 | For all outcomes, present, for each study: (a) summary statistics for each group (where appropriate) and (b) an effect estimate and its precision (e.g. confidence/credible interval), ideally using structured tables or plots. | Figure 3, 4, 5, 6, 7 |
| Results of syntheses | 20a | For each synthesis, briefly summarise the characteristics and risk of bias among contributing studies. | 10 |
| | 20b | Present results of all statistical syntheses conducted. If meta-analysis was done, present for each the summary estimate and its precision (e.g. confidence/credible interval) and measures of statistical heterogeneity. If comparing groups, describe the direction of the effect. | Figure 3, 4, 5, 6, 7, Table S4, S5, S6, S7, Figure S1, S2, S3, S4, S6, S7, S8, S9, S10, S12, S13 |
| | 20c | Present results of all investigations of possible causes of heterogeneity among study results. | Table S4, S5, S6, S7, Figure S1, S2, S3, S4, S6, S7, S8, S9, S10, S12, S13, supplementary sensitivity analyses |
| | 20d | Present results of all sensitivity analyses conducted to assess the robustness of the synthesized results. | supplementary sensitivity analyses |
| Reporting biases | 21 | Present assessments of risk of bias due to missing results (arising from reporting biases) for each synthesis assessed. | Figure S5, S11 |
| Certainty of evidence | 22 | Present assessments of certainty (or confidence) in the body of evidence for each outcome assessed. | 10 |
| **DISCUSSION** | | | |
| Discussion | 23a | Provide a general interpretation of the results in the context of other evidence. | 12 |
| | 23b | Discuss any limitations of the evidence included in the review. | 15 |
| | 23c | Discuss any limitations of the review processes used. | 15 |
| | 23d | Discuss implications of the results for practice, policy, and future research. | 15 |

### PRISMA 2020 Checklist

| Section and Topic | Item # | Checklist item | Location where item is reported |
|---|---|---|---|
| **OTHER INFORMATION** | | | |
| Registration and protocol | 24a | Provide registration information for the review, including register name and registration number, or state that the review was not registered. | 4 |
| | 24b | Indicate where the review protocol can be accessed, or state that a protocol was not prepared. | NA |
| | 24c | Describe and explain any amendments to information provided at registration or in the protocol. | NA |
| Support | 25 | Describe sources of financial or non-financial support for the review, and the role of the funders or sponsors in the review. | 11 |
| Competing interests | 26 | Declare any competing interests of review authors. | 11 |
| Availability of data, code and other materials | 27 | Report which of the following are publicly available and where they can be found: template data collection forms; data extracted from included studies; data used for all analyses; analytic code; any other materials used in the review. | Supplementary material |

*From:* Page MJ, McKenzie JE, Bossuyt PM, Boutron I, Hoffmann TC, Mulrow CD, et al. The PRISMA 2020 statement: an updated guideline for reporting systematic reviews. BMJ 2021;372:n71. doi: 10.1136/bmj.n71

**Table S3.** Search strategy

| PUBMED | Results |
|---|---|
| (“Virtual Reality Exposure Therapy”[MeSH Terms] OR "virtual reality"[MeSH Terms] OR "reality" [Title/Abstract] OR "virtual realities" [Title/Abstract] OR "VR"[Title/Abstract] OR "exergaming"[MeSH Terms] OR "gamification"[MeSH Terms] OR "computer simulation"[MeSH Terms] OR "virtual environment"[Title/Abstract] OR "interactive environment"[Title/Abstract] OR "immersive"[Title/Abstract] OR "Haptic Technology"[MeSH Terms] OR "computerized cognitive training"[Title/Abstract]) **AND** ("Stroke"[MeSH Terms] OR "Stroke"[Title/Abstract] OR "brain vascular accident"[Title/Abstract] OR "brain infarction"[Title/Abstract] OR "cerebral infarction"[Title/Abstract] OR "brain haemorrhage"[Title/Abstract]) **AND** ("Intervention*"[Title/Abstract] OR "program*"[Title/Abstract] OR "trial*"[Title/Abstract] OR "rehabilitation"[Title/Abstract]) | 1781 |
| **SCOPUS** | |
| **TITLE-ABS** ("Virtual Reality" OR reality OR "virtual realities" OR VR OR exergaming OR "active-video gaming" OR gamification OR "computer simulation" OR "Immersion Therapy" OR "Virtual Reality Exposure Therapy" OR "virtual environment" OR "interactive environment" OR immersive) **AND TITLE-ABS** (Stroke OR "Brain Vascular Accident" OR " brain infarction" OR "cerebral infarction" OR " brain haemorrhage") **AND TITLE-ABS** ("Intervention*" OR "program*" OR "trial*" OR "rehabilitation") | 2574 |
| **WOS** | |
| **AB=**("Virtual Reality" OR reality OR "virtual realities" OR VR OR exergaming OR "active-video gaming" OR gamification OR "computer simulation" OR "Immersion Therapy" OR "Virtual Reality Exposure Therapy" OR "virtual environment" OR "interactive environment" OR immersive) **AND AB=**(Stroke OR "Brain Vascular Accident" OR " brain infarction" OR "cerebral infarction" OR " brain haemorrhage") **AND AB=** ("Intervention*" OR "program*" OR "trial*" OR "rehabilitation") | 2415 |
| **COCHRANE** | |
| **Title Abstract Keyword** ("Virtual Reality" OR reality OR "virtual realities" OR VR OR exergaming OR "active-video gaming" OR gamification OR "computer simulation" OR "Immersion Therapy" OR "Virtual Reality Exposure Therapy" OR "virtual environment" OR "interactive environment" OR immersive) AND (Stroke OR "Brain Vascular Accident" OR " brain infarction" OR "cerebral infarction" OR " brain haemorrhage") AND ("Intervention" OR "program" OR "trial" OR "rehabilitation") | 1162 |

**Table S4.** Meta-regression models assessing potential moderators of the effect of virtual reality-based interventions on upper-limb function measured by the Fugl-Meyer Assessment-Upper Extremity (FM-UE).

| Moderator | *β* | SE | 95% CI | *p*-value | $R^2$ (%) | $I^2$ (%) |
|---|---|---|---|---|---|---|
| Time difference between groups | 0.002 | 0.019 | -0.039 to 0.043 | 0.923 | 0 | 84.83 |
| Total duration (weeks) | -0.810 | 0.860 | -2.643 to 1.023 | 0.361 | 0 | 84.71 |
| Total duration (min) | 0.001 | 0.001 | -0.001 to 0.004 | 0.344 | 14.86 | 78.73 |
| Age | -0.052 | 0.048 | -0.154 to 0.050 | 0.294 | 0 | 84.74 |
| Women percentage | -0.036 | 0.061 | -0.166 to 0.095 | 0.571 | 3.12 | 82.41 |

**Legend:** *β* = unstandardized regression coefficient; SE = standard error; 95% CI = 95% confidence interval; *p*-value = significance of the moderator effect; $R^2$ = proportion of variance explained by the moderator; $I^2$ = proportion of residual heterogeneity.

**Table S5.** Meta-regression models assessing potential moderators of the effect of virtual reality-based interventions on upper-limb function measured by the Action Research Arm Test (ARAT).

| Moderator | *β* | SE | 95% CI | *p*-value | $R^2$ (%) | $I^2$ (%) |
|---|---|---|---|---|---|---|
| Time difference between groups | -0.03 | 2.73 | -0.46 to 0.40 | 0.871 | 0 | 97.35 |
| Total duration (weeks) | -2.156 | 0.959 | -4.501 to 0.190 | 0.066 | 59.09 | 90.84 |
| Total duration (min) | -0.001 | 0.003 | -0.008 to 0.007 | 0.826 | 0 | 96.46 |
| Age | 0.180 | 0.388 | -0.819 to 1.778 | 0.663 | 0 | 94.75 |
| Women percentage | -0.398 | 0.191 | -0.889 to 0.093 | 0.091 | 48.48 | 87.23 |

**Legend:** *β* = unstandardized regression coefficient; SE = standard error; 95% CI = 95% confidence interval; *p*-value = significance of the moderator effect; $R^2$ = proportion of variance explained by the moderator; $I^2$ = proportion of residual heterogeneity.

**Table S6.** Meta-regression models assessing potential moderators of the effect of virtual reality-based interventions on upper-limb function measured by the Box and block test (BBT).

| Moderator | *β* | SE | 95% CI | *p*-value | $R^2$ (%) | $I^2$ (%) |
|---|---|---|---|---|---|---|
| Time difference between groups | 0.042 | 1.204 | -3.549 to 2.005 | **0.006** | 78.09 | 47.96 |
| Total duration (weeks) | 0.247 | 1.125 | -2.348 to 2.843 | 0.832 | 0 | 82.42 |
| Total duration (min) | 0.002 | 0.004 | -0.007 to 0.011 | 0.599 | 0 | 81.36 |
| Age | -0.068 | 0.405 | -1.000 to 0.865 | 0.871 | 0 | 82.75 |
| Women percentage | 0.075 | 0.188 | -0.359 to 0.510 | 0.700 | 0 | 81.80 |

**Legend:** *β* = unstandardized regression coefficient; SE = standard error; 95% CI = 95% confidence interval; *p*-value = significance of the moderator effect; $R^2$ = proportion of variance explained by the moderator; $I^2$ = proportion of residual heterogeneity.

**Table S7.** Meta-regression models assessing potential moderators of the effect of virtual reality-based interventions on handgrip strength.

| Moderator | *β* | SE | 95% CI | *p*-value | $R^2$ (%) | $I^2$ (%) |
|---|---|---|---|---|---|---|
| Time difference between groups | 0.192 | 0.101 | -0.088 to 0.473 | 0.129 | 14.18 | 95.52 |
| Total duration (weeks) | -0.022 | 0.916 | -2.565 to 2.520 | 0.982 | 0 | 92.18 |
| Total duration (min) | -0.001 | 0.002 | -0.007 to 0.005 | 0.745 | 0 | 91.84 |
| Age | -0.814 | 0.170 | -1.355 to -0.272 | **0.017** | 90.98 | 45.75 |
| Women percentage | -0.327 | 0.183 | -1.113 to 0.460 | 0.216 | 46.92 | 86.88 |

**Legend**: *β* = unstandardized regression coefficient; SE = standard error; 95% CI = 95% confidence interval; *p*-value = significance of the moderator effect; $R^2$ = proportion of variance explained by the moderator; $I^2$ = proportion of residual heterogeneity.

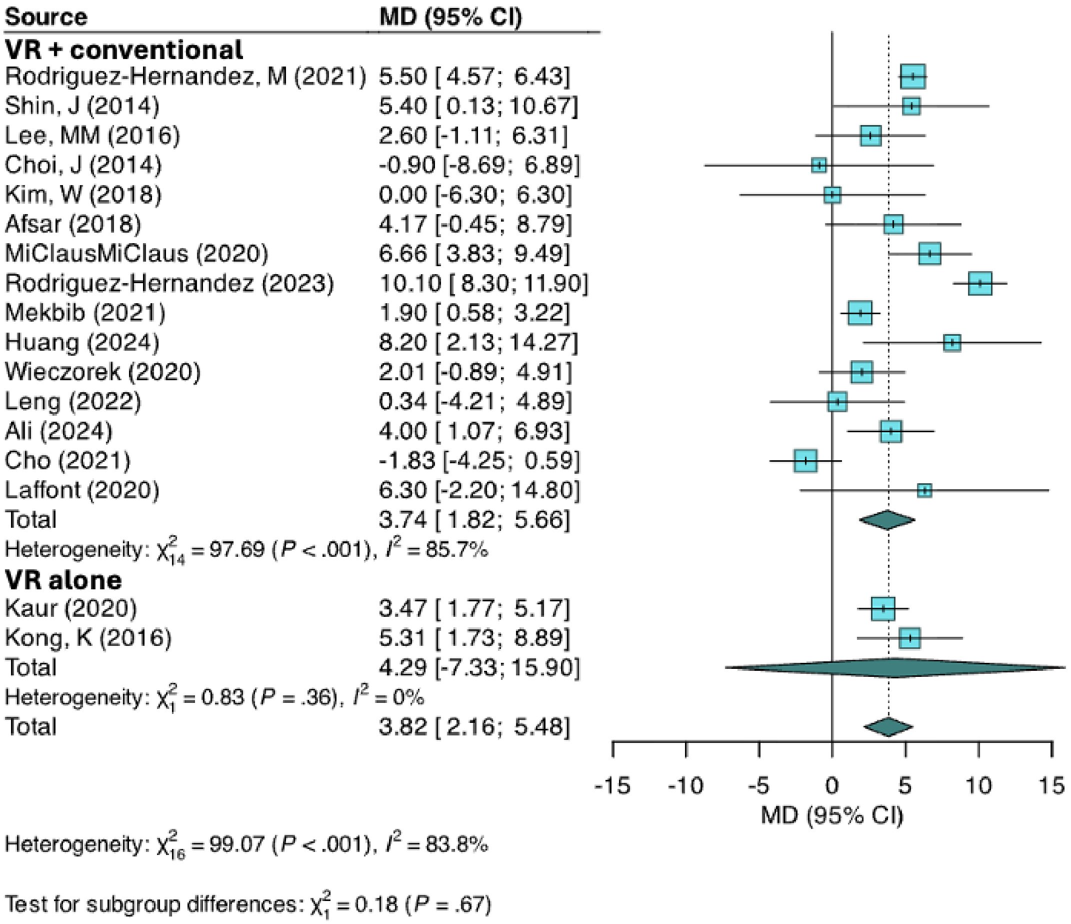


**Legend**: VR = virtual reality; MD = mean difference; 95% CI = 95% confidence interval.

**Figure S1.** Forest plot of subgroup analysis comparing virtual reality interventions alone versus virtual reality combined with conventional therapy on upper-limb function measured with the Fugl-Meyer Assessment-Upper Extremity (FM-UE).

| Source | MD (95% CI) |
|---|---|
| **Commercial** | |
| Rodriguez-Hernandez, M (2021) | 5.50 [ 4.57; 6.43] |
| Lee, MM (2016) | 2.60 [-1.11; 6.31] |
| Choi, J (2014) | -0.90 [-8.69; 6.89] |
| Kaur (2020) | 3.47 [ 1.77; 5.17] |
| Afsar (2018) | 4.17 [-0.45; 8.79] |
| Huang (2024) | 8.20 [ 2.13; 14.27] |
| Leng (2022) | 0.34 [-4.21; 4.89] |
| Cho (2021) | -1.83 [-4.25; 0.59] |
| Kong, K (2016) | 5.31 [ 1.73; 8.89] |
| Total | 3.05 [ 0.65; 5.45] |
| Heterogeneity: $\chi^2_8 = 39.26$ ($P < .001$), $I^2 = 79.6\%$ | |
| **Specialised** | |
| Shin, J (2014) | 5.40 [ 0.13; 10.67] |
| Kim, W (2018) | 0.00 [-6.30; 6.30] |
| MiClausMiClaus (2020) | 6.66 [ 3.83; 9.49] |
| Rodriguez-Hernandez (2023) | 10.10 [ 8.30; 11.90] |
| Mekbib (2021) | 1.90 [ 0.58; 3.22] |
| Wieczorek (2020) | 2.01 [-0.89; 4.91] |
| Ali (2024) | 4.00 [ 1.07; 6.93] |
| Laffont (2020) | 6.30 [-2.20; 14.80] |
| Total | 4.66 [ 1.88; 7.45] |
| Heterogeneity: $\chi^2_7 = 59.43$ ($P < .001$), $I^2 = 88.2\%$ | |
| Total | 3.82 [ 2.16; 5.48] |

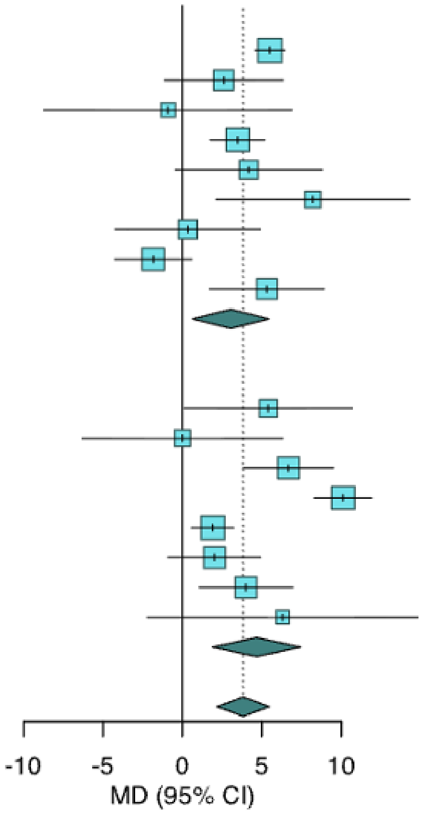


Heterogeneity: $\chi^2_{16} = 99.07$ ($P < .001$), $I^2 = 83.8\%$

Test for subgroup differences: $\chi^2_1 = 1.06$ ($P = .30$)

**Legend**: MD = mean difference; 95% CI = 95% confidence interval.

**Figure S2.** Forest plot of subgroup analysis comparing commercial versus specialised virtual reality virtual reality system for upper-limb function measured with the Fugl-Meyer Assessment-Upper Extremity (FM-UE).

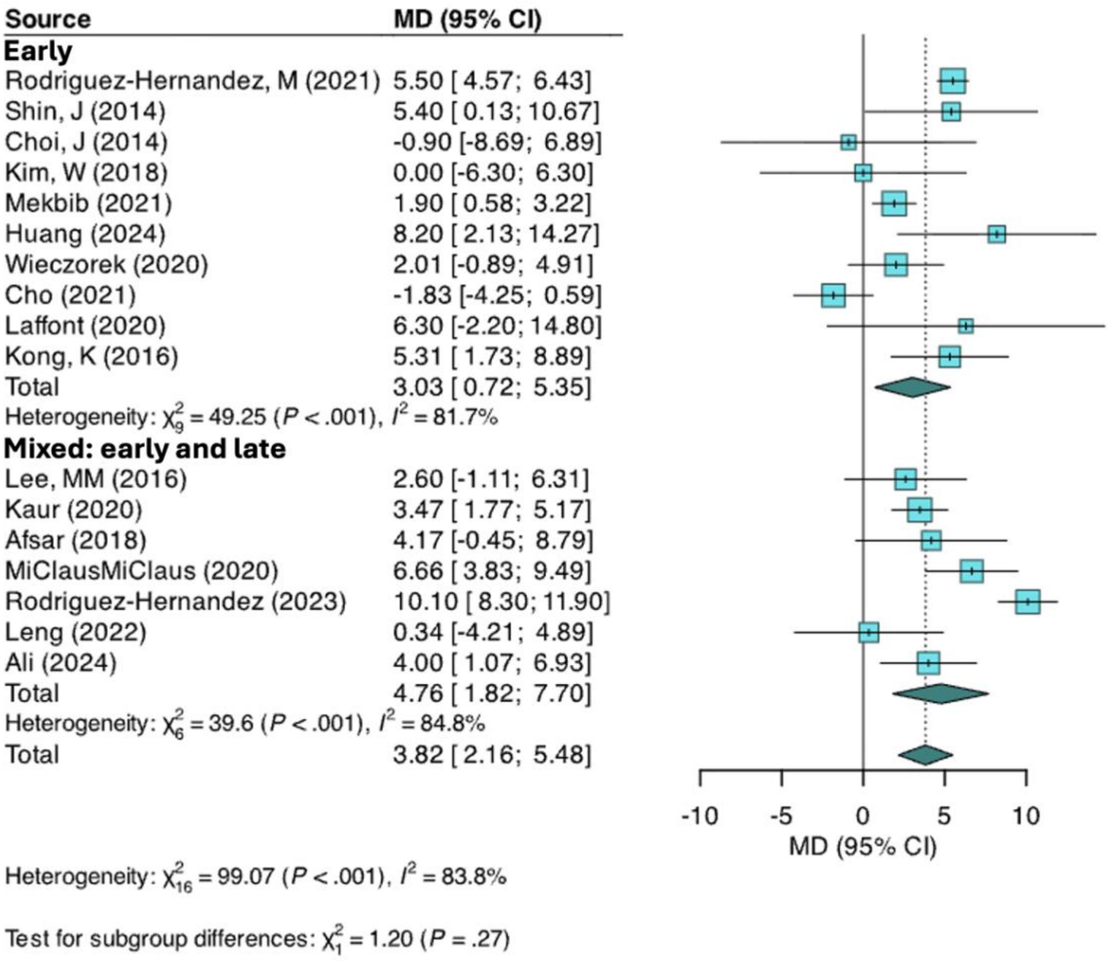


**Legend**: MD = mean difference; 95% CI = 95% confidence interval.

**Figure S3.** Forest plot of subgroup analysis comparing studies with early-phase patients only versus studies with mixed early and late-phase patients for upper-limb function measured with the Fugl-Meyer Assessment-Upper Extremity (FM-UE).

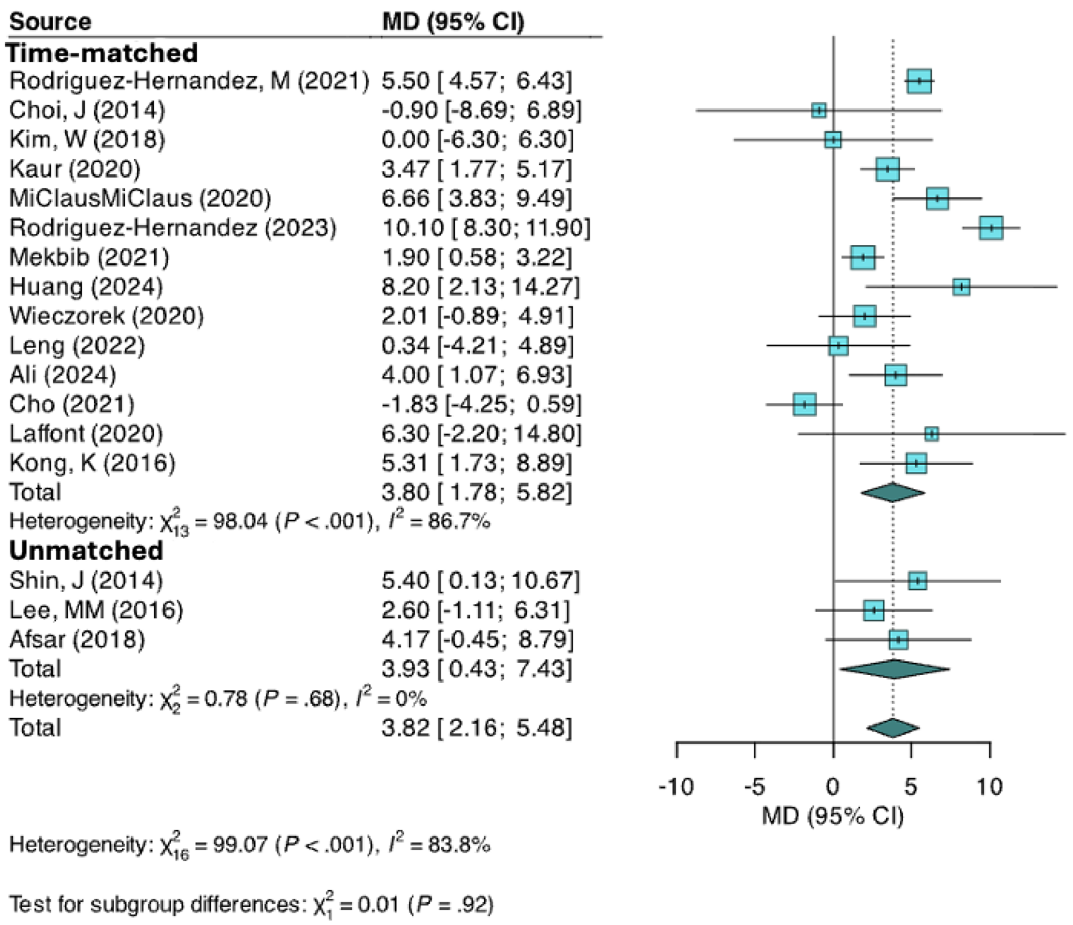


**Legend**: MD = mean difference; 95% CI = 95% confidence interval.

**Figure S4.** Forest plot of subgroup analysis comparing time-matched versus unmatched intervention durations in virtual reality studies on upper-limb function measured with the Fugl-Meyer Assessment-Upper Extremity (FM-UE).

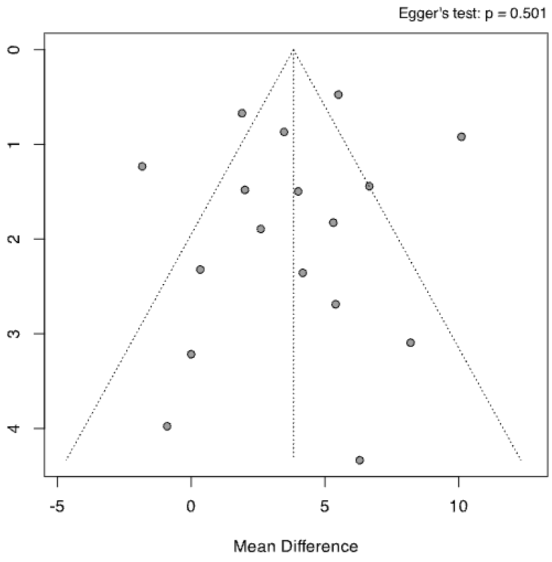


**Figure S5.** Funnel plot assessing publication bias for studies reporting Fugl-Meyer Assessment-Upper Extremity (FM-UE).

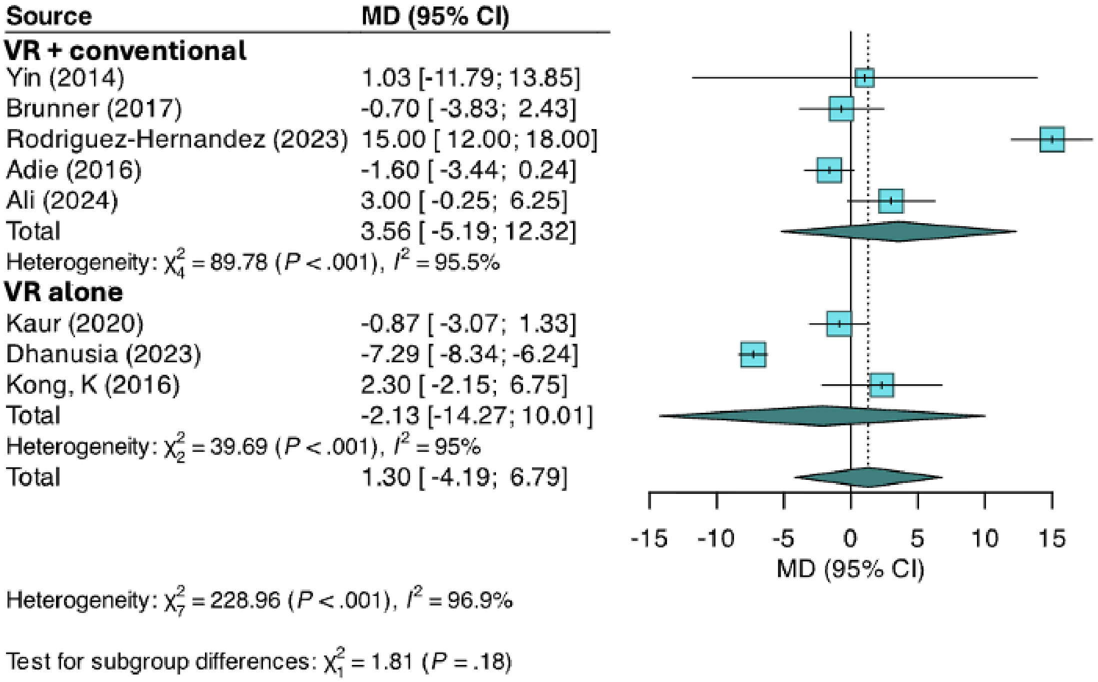


**Figure S6.** Forest plot of subgroup analysis comparing virtual reality interventions alone versus virtual reality combined with conventional therapy on upper-limb function measured with the Action Research Arm Test (ARAT).

**Legend**: VR = virtual reality; MD = mean difference; 95% CI = 95% confidence interval.

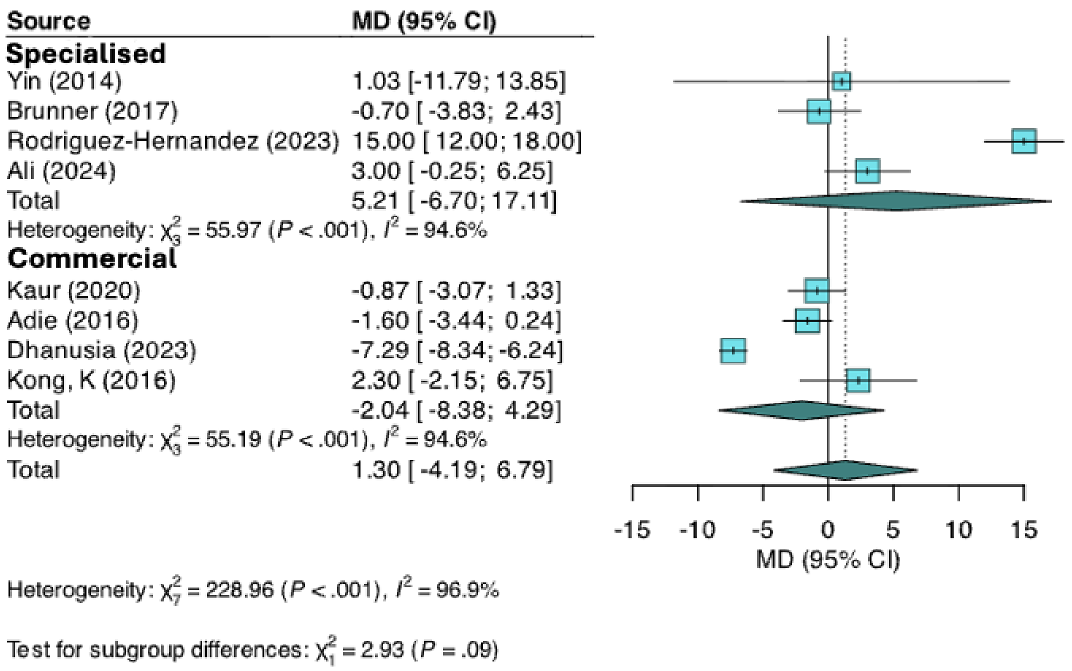


**Figure S7.** Forest plot of subgroup analysis comparing specialised versus commercial virtual reality virtual reality system for upper-limb function measured with the Action Research Arm Test (ARAT)

**Legend**: MD = mean difference; 95% CI = 95% confidence interval.

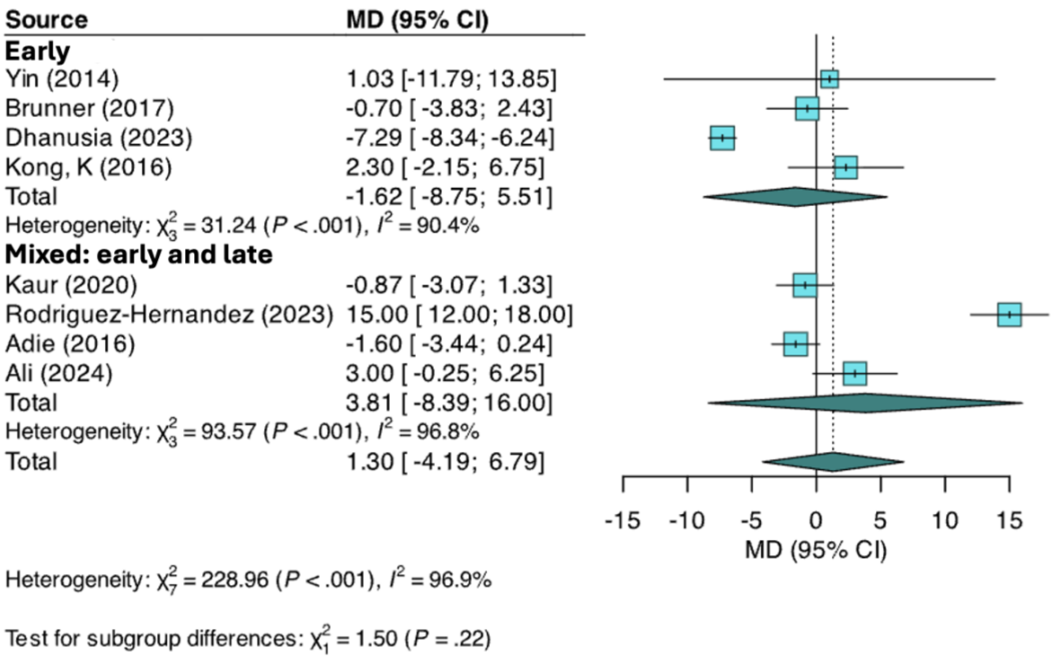


**Figure S8.** Forest plot of subgroup analysis comparing studies with early-phase patients only versus studies with mixed early and late-phase patients for upper-limb function measured with the Action Research Arm Test (ARAT).

**Legend**: MD = mean difference; 95% CI = 95% confidence interval

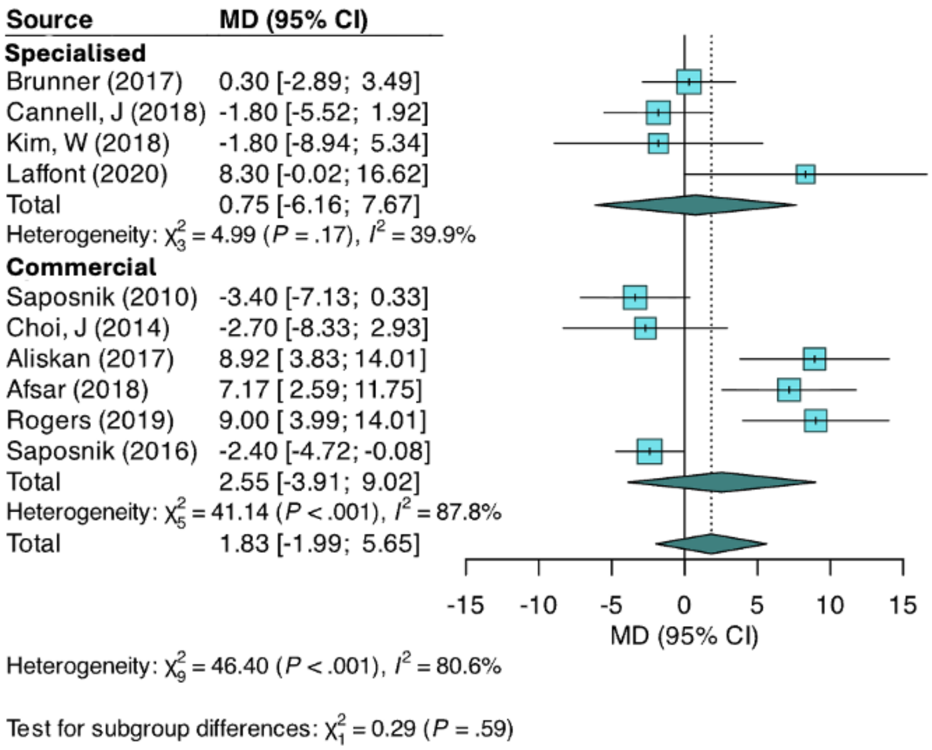


**Figure S9.** Forest plot of subgroup analysis comparing specialised versus commercial virtual reality virtual reality system for upper-limb function measured with the Box and Block Test (BBT).

**Legend**: MD = mean difference; 95% CI = 95% confidence interval

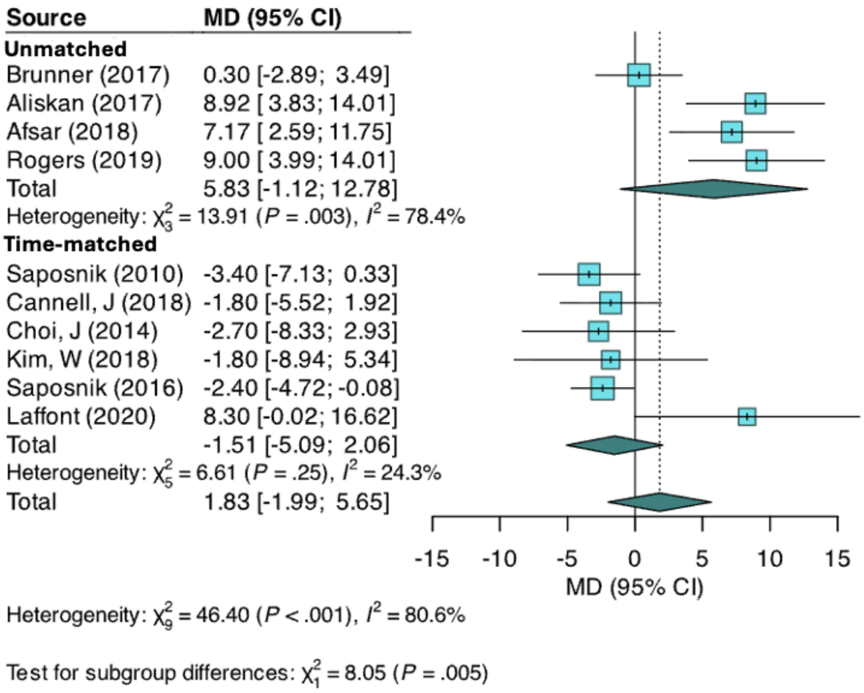


**Figure S10.** Forest plot of subgroup analysis comparing time-matched versus unmatched intervention durations in virtual reality studies on upper-limb function measured with the Box and Block Test (BBT).

**Legend**: MD = mean difference; 95% CI = 95% confidence interval

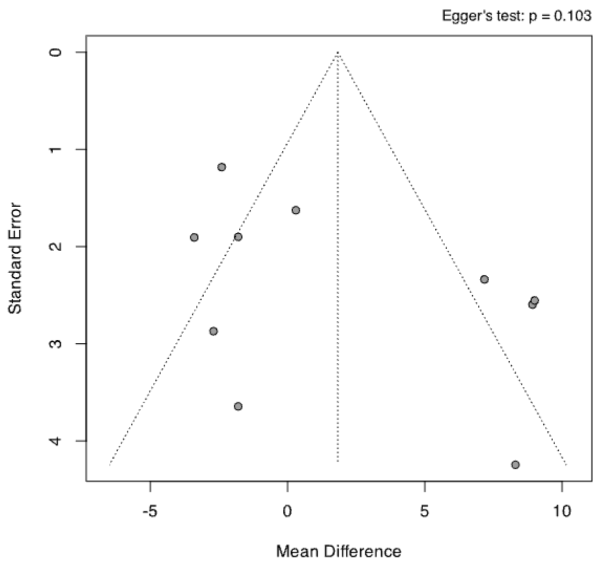


**Figure S11.** Funnel plot assessing publication bias for studies reporting Box and Block Test (BBT).

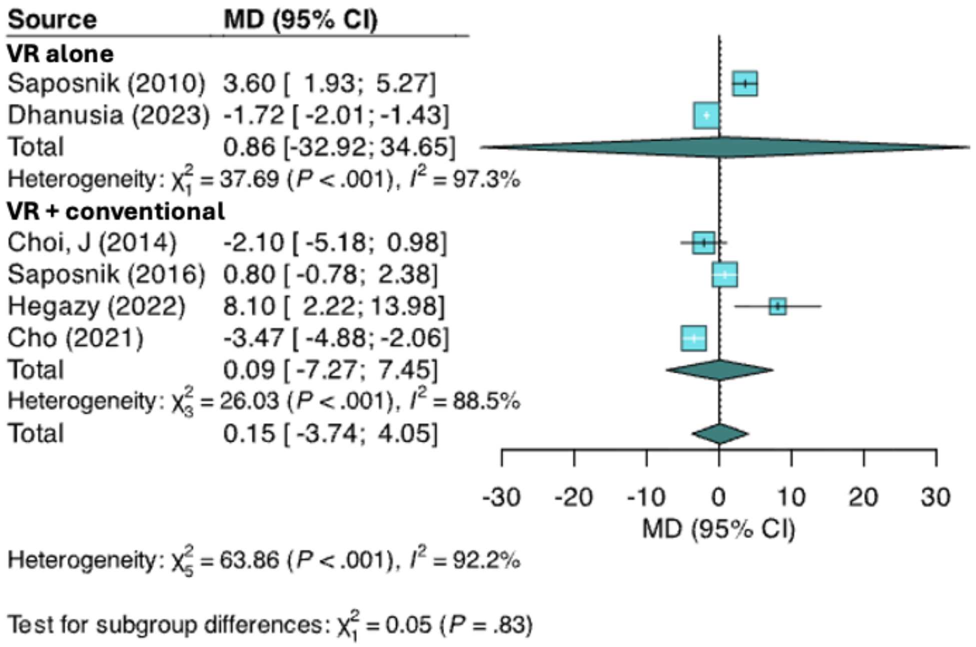


**Legend**: VR = virtual reality; MD = mean difference; 95% CI = 95% confidence interval.

**Figure S12.** Forest plot of subgroup analysis comparing virtual reality interventions alone versus virtual reality combined with conventional therapy on handgrip strength.

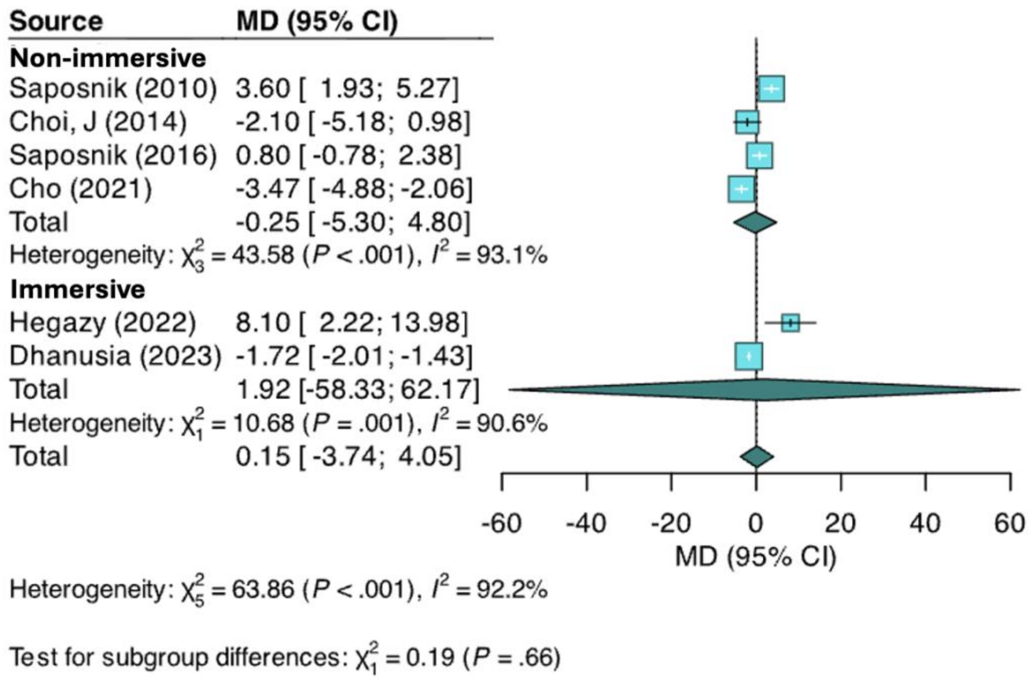


**Legend**: MD = mean difference; 95% CI = 95% confidence interval.

**Figure S13.** Forest plot of subgroup analysis comparing immersive versus non-immersive virtual reality interventions on handgrip strength.

Subgroup/meta-regression analyses/publication bias assessment not shown were not feasible due to insufficient data.
